# A novel two-sample Mendelian randomization framework integrating common and rare variants: application to assess the effect of HDL-C on preeclampsia risk

**DOI:** 10.1101/2025.08.20.25334100

**Authors:** Yu Zhang, Ming Li, David M. Haas, C. Noel Bairey Merz, Tsegaselassie Workalemahu, Kelli Ryckman, Janet M. Catov, Lisa D. Levine, Alexa Freedman, George R. Saade, Jiaqi Hu, Hongyu Zhao, Xihao Li, Nianjun Liu, Qi Yan

**Affiliations:** Department of Epidemiology and Biostatistics, Indiana University School of Public Health-Bloomington, Bloomington, IN 47405, USA; Department of Obstetrics and Gynecology, Indiana University School of Medicine, Indianapolis, IN 46202, USA; Barbra Streisand Women’s Heart Center, Smidt Heart Institute, Cedars-Sinai Medical Center, Los Angeles, CA 90048, USA; Department of Obstetrics and Gynecology, University of Utah Health, Salt Lake City, UT 84112 USA; Departments of Obstetrics, Gynecology and Reproductive Sciences and the Department of Epidemiology, University of Pittsburgh, PA 15213, USA; Department of Obstetrics and Gynecology, The University of Pennsylvania Perelman School of Medicine, Philadelphia, PA 19104, USA; Department of Preventive Medicine, Northwestern University Feinberg School of Medicine, Chicago, IL 60611, USA; Department of Obstetrics and Gynecology, Eastern Virginia Medical School, Norfolk, VA 23507, USA; Department of Chronic Disease Epidemiology, Yale School of Public Health, New Haven, CT 06510, USA; Department of Biostatistics, School of Public Health, Yale University, New Haven, CT 06520, USA; Department of Biostatistics, University of North Carolina at Chapel Hill, Chapel Hill, NC 27599, USA; Department of Genetics, University of North Carolina at Chapel Hill, Chapel Hill, NC 27599, USA; Department of Obstetrics and Gynecology, Columbia University, New York, NY 10032, USA

**Keywords:** Mendelian randomization, rare variants, common variants, STAARpipeline, lipids, preeclampsia

## Abstract

Mendelian randomization (MR) has become an important technique for establishing causal relationships between risk factors and health outcomes. By using genetic variants as instrumental variables, it can mitigate bias due to confounding and reverse causation in observational studies. Current MR analyses have predominantly used common genetic variants as instruments, which represent only part of the genetic architecture of complex traits. Rare variants, which can have larger effect sizes and provide unique biological insights, have been understudied due to statistical and methodological challenges. We introduce MR-CARV, a novel framework integrating common and rare genetic variants in two-sample Mendelian randomization. This method leverages comprehensive genetic data made available by high-throughput sequencing technologies and large-scale consortia. Rare variants are aggregated into functional categories, such as gene-coding, gene-noncoding, and non-gene regions, by leveraging variant annotations and biological impact as weights. The effects of rare variant sets are then estimated with STAARpipeline and combined with the estimated effects of common variants by the existing MR methods. Simulation studies demonstrate that MR-CARV maintains robust type I error and achieves higher statistical power, with up to a 66.3% relative increase compared to existing methods only based on common variants. Consistent with these findings, application to real data on HDL-C and preeclampsia showed that MR-CARV(IVW) yielded a more precise and statistically significant effect estimate (–0.020, SE = 0.0102, P = 0.0470) than IVW using only common variants (–0.023, SE = 0.0123, P = 0.0659).

## 1 Introduction

Mendelian randomization (MR) has emerged as a powerful tool for inferring causal relationships between risk factors and health outcomes, leveraging genetic variants as instrumental variables to circumvent the inherent issues of confounding and reverse causation in observational studies [1, 2]. Traditionally, MR analyses have primarily utilized common genetic variants as instruments, which account for a limited spectrum of the genetic architecture underlying complex diseases and traits. Rare variants, despite their relatively low minor allele frequencies (MAF) (less than 1% or 5%), may have larger effect sizes and provide insights into novel biological mechanisms and pathways [3, 4]. But they have been largely overlooked in MR.

Integrating rare variants into two-sample MR analysis has been limited by several key challenges. First, individual rare variants contribute minimal variation in allele frequency across the population, leading to reduced statistical power in genome-wide association studies (GWAS) to detect associations with exposures [5]. Second, even when rare variants exhibit moderate associations with exposures, they often fail to pass stringent genome-wide significance thresholds, leading to their exclusion during instrument selection in conventional MR pipelines. Third, rare variants typically require aggregation strategies—such as burden tests or kernel methods—to capture their collective effects [6–8], which are not directly compatible with standard MR techniques that rely on independent, variant-level effect estimates. As a result, rare variants are rarely selected as instruments in two-sample MR studies [9–12], limiting the statistical power of MR analysis by overlooking an important aspect of the genetic architecture.

The advent of high-throughput whole-genome sequencing (WGS) technologies [13] and largescale genomic consortia [14] has enabled the generation of extensive genetic data, including rare variants. These developments have driven the need for scalable statistical tools capable of leveraging rare variant information from WGS data. For instance, STAARpipeline [15] performs genome-wide rare variant association tests using gene-based burden [6] or kernel methods [7, 8] while incorporating functional annotations and controlling for population structure. As a result, functionally informed burden-level summary statistics for rare variants are becoming increasingly available. However, despite these advances, current MR methods are not designed to integrate burden-level rare variant statistics, as they typically rely on SNP-level GWAS summary data.

In this study, we introduce a novel method called, two-sample MR framework that integrates both Common and Annotation-informed Rare Variants, referred to as MR-CARV. By using both common and rare variants as instruments, MR-CARV addresses limitations of existing MR methods, and maximizes the utility of genetic data. Our simulation studies demonstrate that this new framework maintains the correct type I error rates and achieves higher statistical power than existing methods only based on common variants. We applied the MR-CARV framework to explore the causal relationship between lipid levels and preeclampsia, given lipid metabolism’s critical role in pregnancy and its links to complications like preeclampsia [16–18]. Although lipid levels is known to be associated with pregnancy complications [19], it has not been well-estimated whether these associations are causal. Our analysis underscores the value of integrating rare variants in MR methods and offers new insights into the biological connection between lipids and preeclampsia.

## 2 Material and methods

Suppose we have *I* common variants as valid instruments for evaluating the causal effect (*θ*_0_) of an exposure variable, *X*, on an outcome variable, *Y* . For the *i*-th common variant, we obtain its estimated effect on the exposure, denoted as 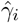, and its standard error, 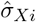 from one study. Similarly, we obtain its estimated effect on the outcome, denoted as 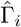, and its standard error, 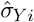 from another study that is independent from the previous one. There are various MR techniques designed to estimate *θ*_0_. In this study, we selected some commonly used methods and extended them using the MR-CARV framework.

### 2.1 Inverse variance weighted (IVW) method

For each common variant, *θ*_0_ is estimated as 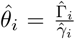. The IVW [20], one of the most widely used MR methods, combines these estimates weighted by the inverse of their variances. Its estimator [20] is given by:

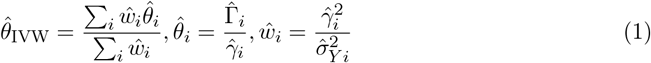

with variance being

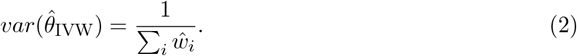

This approach assumes that the genetic variants are valid instruments, namely, they are associated with the exposure, influence the outcome only through the exposure, and are not associated with any confounders of the exposure-outcome relationship.

In the following we describe our proposed strategy to further integrate rare variants into MR-CARV framework. Suppose there are *J* rare variant sets and the *j*-th rare variant set has *p*_*j*_ rare variants. We combine rare variants within each set into a Burden variable using a weighted method as employed in STAAR [21]. We start by identifying *K* functional annotations for rare variants, defined as variants with MAF less than 1% or 5%. These annotations provide insights into the biological relevance and potential impact of each variant, guiding the weighting process. The Burden variable, *B*_*jk*_, for the *j*-th rare variant set with the *k*-th annotation is constructed by weighting the genotype of each rare variant within the set, *G*_*jl*_, *l* = 1, 2, …, *p*_*j*_, with the product of the functional annotation, *π*., and a weighting based on the allele frequency, *λ*. = *Beta*(MAF., *a*_1_, *a*_2_). As such, 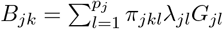. Following STAAR, we set (*a*_1_, *a*_2_) to (1, 25) to upweight rarer variants, and to (1, 1) for equal weighting of all rare variants. We also set *π*_*jkl*_ = 1 for *k* = 0 to only include the *λ*. as the weight. With the Burden variable representing the aggregated effect of rare variants in each set, we conduct rare variant association study (RVAS) to estimate the genetic effect of each rare variant set on the exposure 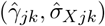 and outcome 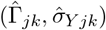, respectively. Note that the *J* Burden variables should satisfy the assumption of IVW to be valid instrument, the same as for the common variants. The RVAS results for *J* rare variant sets with the *k*-th annotation along with common variants are then incorporated into equation (1) to get the IVW estimator of the proposed framework (MR-CARV(IVW)) that considers both common and rare variants, which is

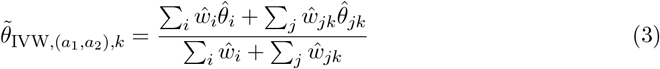

with variance being

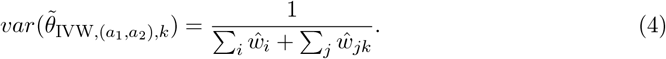

Similar to 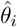 and 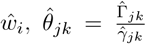 and 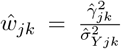. The P-value of 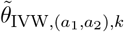 is denoted by 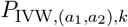. We then combine the P-values 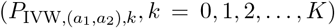 under *K* annotations and different settings of (*a*_1_, *a*_2_) into one P-value using the Cauchy combination test [22]. The combined statistics is

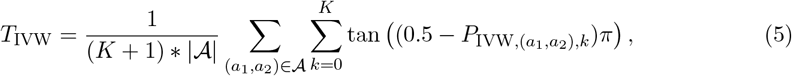

where the 𝒜 is the set of specified values of (*a*_1_, *a*_2_), and |𝒜 | is the size of set 𝒜. We set 𝒜 = {(1, 25), (1, 1)} in practice as in STAAR [21]. The P-value of MR-CARV(IVW) can be approximated by

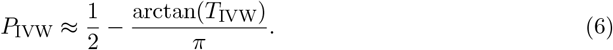

### 2.2 Debiased inverse variance weighted (dIVW) method

However, the IVW method is subject to several sources of bias, which can compromise the validity of causal inference. One significant source of bias in IVW estimation arises from weak instruments—genetic variants that have weak associations with the exposure. In such cases, the estimated SNP-exposure effects 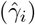 are small and imprecise, leaning to unstable ratio estimates 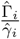 and inflated variance. The dIVW [5] addresses this issue with a bias correction factor. The dIVW estimator [5] for common variants is given by

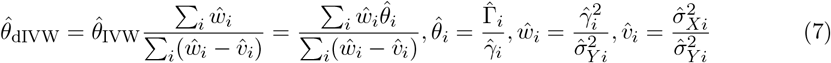

It’s variance [5] is

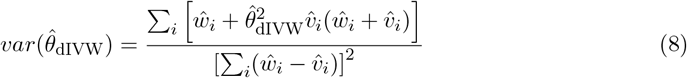

Similar to MR-CARV(IVW), we construct MR-CARV(dIVW) by incorporating the summary statistics of rare variant sets with *k*-th annotation and the parameter combination (*a*_1_, *a*_2_). The formula would be

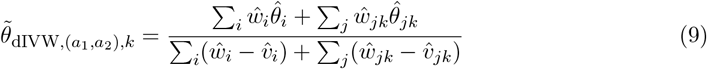

with variance being

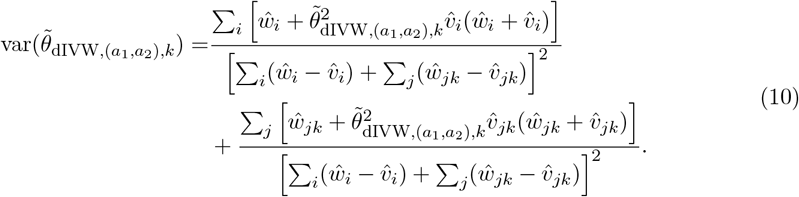

We then combine the P-values of MR-CARV(dIVW) under all the annotations and settings of (*a*_1_, *a*_2_) using the Cauchy combination test as in MR-CARV(IVW).

### 2.3 Robust adjusted profile score (RAPS) method

Another important source of bias in IVW estimation is horizontal pleiotropy, which occurs when genetic instruments influence the outcome through pathways other than the exposure. This violates the exclusion restriction assumption in MR and can lead to biased causal effect estimates, particularly when pleiotropic effects are heterogeneous. The RAPS method [23] addresses pleiotropy by modeling it explicitly through a random effects framework. It assumes that the estimated effects of instruments on the outcome 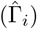 follow a normal distribution with mean *γ*_*i*_*θ*_0_ and variance 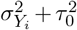, where 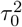 captures the overdispersion due to pleiotropy. Meanwhile, the estimated effects on the exposure are modeled as 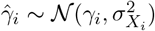. The causal effect is estimated by maximizing the adjusted profile likelihood, and robustness is enhanced by replacing the standard *l*2-loss with alternative loss functions (Huber or Tukey’s biweight loss) to mitigate the influence of outlier SNPs with large pleiotropic effects—referred to as idiosyncratic pleiotropy. To extend RAPS for rare variant analysis in our framework MR-CARV(RAPS), we include the summary statistics of rare variant sets under *k*-th annotation and the parameter combination (*a*_1_, *a*_2_) into the random effects models. Specifically, for common variants, we have 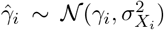 and 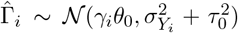, and for rare variant sets, we have 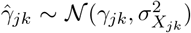 and 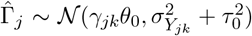 for the Burden variables. After obtaining the P-values under all the annotation and settings of (*a*_1_, *a*_2_), we still use the Cauchy combination test to calculate the P-value of MR-CARV(RAPS). In addition to IVW, dIVW, RAPS, this framework is flexible and can be extended to other MR methods in a similar way as described above. In order to make causal inference, both common variants and Burden variables constructed from rare variants should satisfy the corresponding model assumptions.

### 2.4 Find the uncorrelated instruments

For the IVW, dIVW, and RAPS methods in MR-CARV framework, independent instruments are essential [24]. To achieve this, independent instruments can be selected through established methods, such as choosing a single variant per locus [25] or linkage disequilibrium (LD) pruning [26]. For LD pruning, burden variables can be created for rare variant sets and pruning can be applied to both common variants and burden variables using the STAARpipeline [15]. Alternatively, an automated approach can use individual-level data to calculate correlations be-tween common variants and Burden variables, followed by a graph-based method [27] to select uncorrelated instruments. Detailed steps with an example are provided in the Supplementary Materials Section 1. This process ensures uncorrelated instruments, meeting the requirements of the IVW, dIVW, and RAPS methods. Individual-level data for this can come from study samples or a representative large reference panel with whole sequencing data, such as the UK Biobank [28] provided they align with the study population.

### 2.5 Simulation setting

To evaluate the performance of the methods that consider either common variants alone or both common and rare variants, we conducted a simulation study using genetic data from the 1000 Genomes Project [29]. To avoid issues caused by LD, we selected one independent instrumental variable per chromosome from chromosomes 15 through 22, designating chromosomes 15–18 for common variants and 19–22 for rare variant sets to assess the contribution of each. Utilizing the R package sim1000G [30], we randomly selected 50 SNPs per chromosome from the 10kb region between 30,000,000 and 30,010,000 base pairs (bp), with MAF spanning from 0.001 to 0.5. For common variants, we randomly selected one SNP per chromosome from chromosomes 15-18 with MAF *>* 0.1. For rare variant sets on chromosomes 19-22, we included all variants with MAFs between 0.001 and 0.05. This approach resulted in four common variants and four rare variant sets.

Using these variants, we generated a dataset comprising *N*_*x*_ individuals with exposure, and *N*_*y*_ individuals with outcome. The exposure *X* of *N*_*x*_ individuals was modeled as a linear function of genotypic data for continuous exposure:

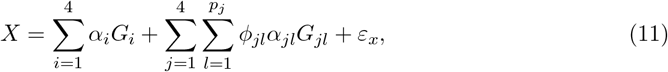

and as an expit function for binary exposure:

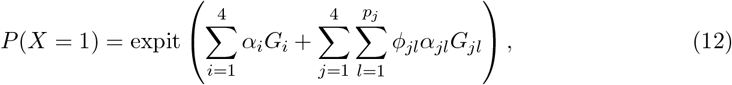

where 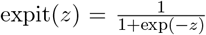. Here, *G*_*i*_, for *i* = 1, 2, 3, 4, was the genotype of the *i*-th common variant, and *G*_*jl*_, for *j* = 1, 2, 3, 4 and *l* = 1, 2, …, *p*_*j*_, denoted the genotype of the *l*-th rare variant in the *j*-th rare variant set. The binary variable *ϕ*_*jl*_ indicated whether a rare variant was causal, determined using a Bernoulli distribution with the probability 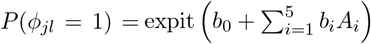, where *A*_*i*_(*i* = 1, 2, …, 5) represented five randomly selected functional annotations from a set of ten. Each of these ten annotations was draw from an independent standard normal distribution (*N* (0, 1)). Here, *b*_*i*_ = log(5) for each selected annotation, and *b*_0_ was set to logit(0.015) or logit(0.18) to achieve approximately 15%, and 35% proportions of causal rare variants (causal variant ratio), respectively [21]. This method ensured that the causality of a rare variant was influenced by a specific set of functional annotations. The effect sizes of these variants on the exposure were represented by *α*_*i*_ for common variants and *α*_*jl*_ for rare variants. In our simulations, *α*_*i*_ was fixed at 0.5 for each common variant. For rare variants, the effect size was determined by *α*_*jl*_ = *c*_0_|log_10_MAF_*jl*_|, where *c*_0_ was set to 0.5. This produced a maximum effect size of 1.5 for variants with MAF = 0.001, and minimum effect size of 0.65 for variants with MAF = 0.05, capturing the influence of MAF on the exposure. Additionally, we assessed the impact of effect direction consistency of rare variants by varying the proportion of rare variants with positive effects in a set on the exposure (i.e., positive effect ratio) to 100%, 80% and 50%. The error term, *ε*_*x*_, encapsulating all random variations, is modeled to follow a standard normal distribution, N(0,1).

To generate *N*_*y*_ individuals with outcome, we first simulated the exposure variable *X* as described previously. For a continuous outcome, *Y* is modeled as a linear function of *X*:

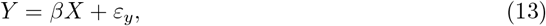

whereas for a binary outcome, the probability of *Y* = 1 is given by

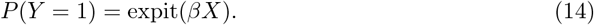

Here, *β* represents the true causal effect of *X* on *Y*, and *ε*_*y*_ is a normally distributed error term following *N* (0, 1).

We conducted simulation studies using large-scale genetic datasets (*N*_*x*_ = *N*_*y*_ = 10,000) to evaluate our framework across both continuous and binary exposure–outcome combinations. Type I error was assessed under the null hypothesis (*β* = 0) across 10,000 iterations, and power was evaluated over 1,000 simulations using a range of positive and negative *β* values (Supplementary Table S1). We generated summary statistics using the STAARpipeline on independent samples and compared existing MR methods (IVW, dIVW, RAPS) with our proposed MR-CARV methods (MR-CARV(IVW), MR-CARV(dIVW), MR-CARV(RAPS)). The MR-CARV methods incorporated MAF weighting (Beta(1, 1) and Beta(1, 25)) and 10 functional annotations. To assess robustness of type I error and power under varying sample sizes, we additionally simulated smaller datasets with combinations of *N*_*x*_ and *N*_*y*_ set to 1,000 and 2,000 for both continuous and binary traits. Further evaluations under LD structure and horizontal pleiotropy were conducted using continuous traits, as representative scenarios to illustrate the core method performance.

## 3 Results

### 3.1 Simulation results

The MR-CARV methods (MR-CARV(IVW), MR-CARV(dIVW), and MR-CARV(RAPS)) ef-fectively controled the type I error rate, comparable to existing methods (IVW, dIVW, and RAPS) under large sample size (**Figure 1**). The type I error rates of IVW, dIVW, MRCARV(IVW) and MR-CARV(dIVW) were consistently below 0.05 across all settings. In contrast, RAPS and MR-CARV(RAPS) maintained type I error rate near 0.05.

**Figure 1.**
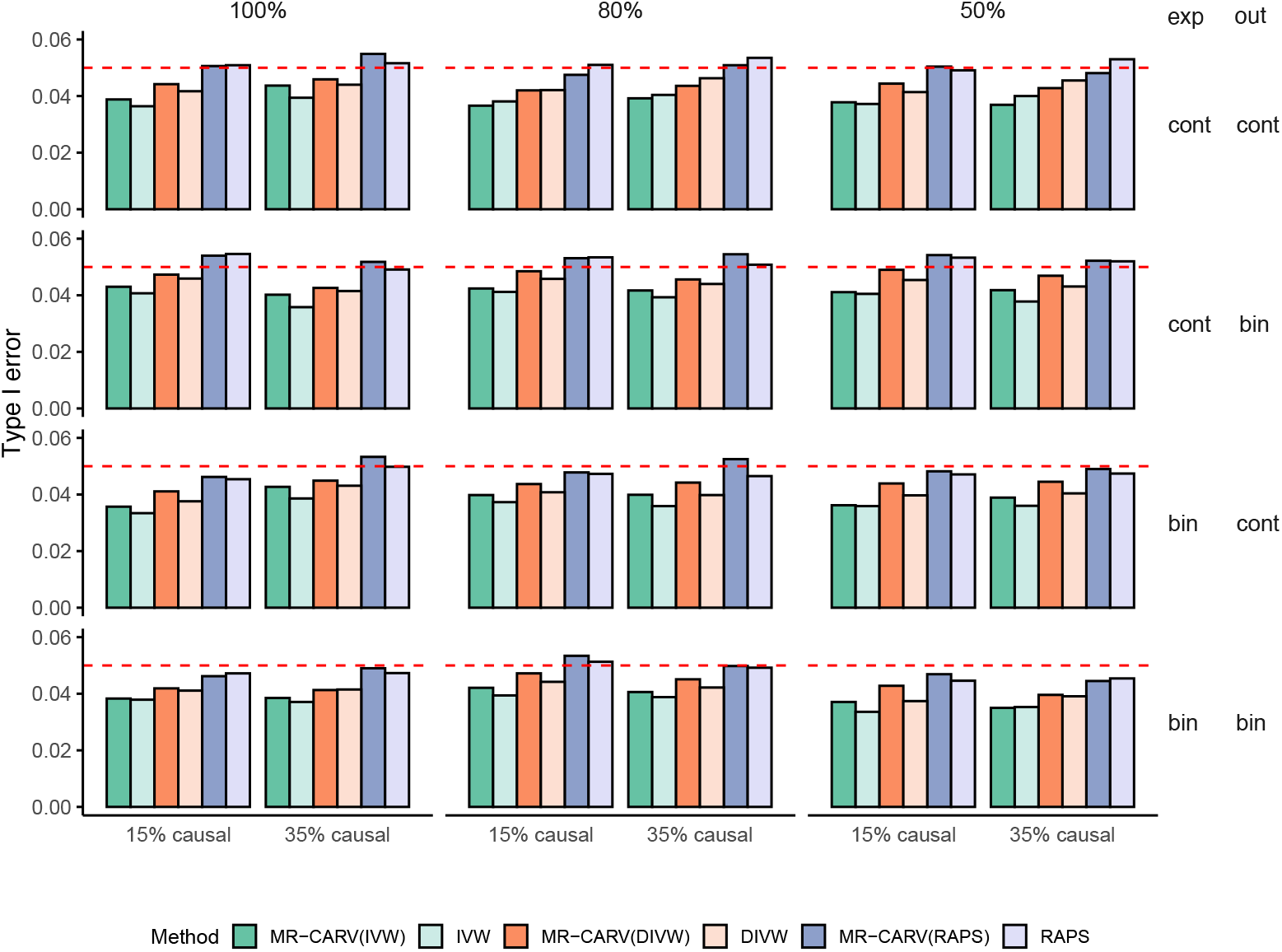
Type I error rate for different methods under various exposure and outcome settings, based on 10,000 simulations with sample size 10,000 for both exposure and outcome (*β* = 0). The first row displays results for continuous exposure and continuous outcome. The second row shows continuous exposure and binary outcome. The third row presents binary exposure and continuous outcome. The fourth row illustrates binary exposure and binary outcome. Each column represents scenarios where 100%, 80%, and 50% of the variants have positive effects on the exposure. The X-axis shows the proportions of causal variants in the signal region, with 15% and 35% causal variants, respectively. The last two columns display the data types for exposure (exp) and outcome (out), respectively (cont: continuous, bin: binary). ALT Text: Bar plots showing type I error rates for different MR methods across scenarios with various combinations of exposure/outcome types, causal variant proportions, and positive effect ratios. Each plot compares MR methods, demonstrating error control consistency when *β* = 0 under simulation conditions.

For both the exposure and outcome were continuous and the true effect size *β* = 0.04, the MR-CARV methods consistently demonstrated higher statistical power compared to the existing methods (first row in **Figure 2**). This superior performance can be attributed to the inclusion of both common and rare variants in the MR-CARV framework, which captures a broader spectrum of genetic variation. By leveraging the additional information provided by rare variants, the MR-CARV methods enhanced the detection of true causal relationships, leading to increased statistical power. This difference in statistical power between MR-CARV methods and existing methods increased with the increase of causal variant ratio and positive effect ratio. Notably, MR-CARV(IVW) achieved the highest power of 95.8% when the causal variant ratio was 35% and the positive effect ratio was 100%, compared to 57.6% with IVW using only common variants—a 66.3% relative increase in power. This is likely because these conditions strengthen the contribution of rare variants, thereby providing even more significant gains in detection capability for the MR-CARV methods. Additionally, the statistical power of MR-CARV methods increased with the increase of the causal variant ratio and the positive effect ratio. In contrast, the statistical power of the existing methods remains relatively stable across different causal ratios and positive ratios because these methods primarily rely on common variants, which are less sensitive to changes in the proportion of causal or positive effect variants. In the scenario of continuous exposure and binary outcome (*β* = 0.06), the performance is similar to that observed in the continuous exposure and continuous outcome setting (the second row in **Figure 2**).

**Figure 2.**
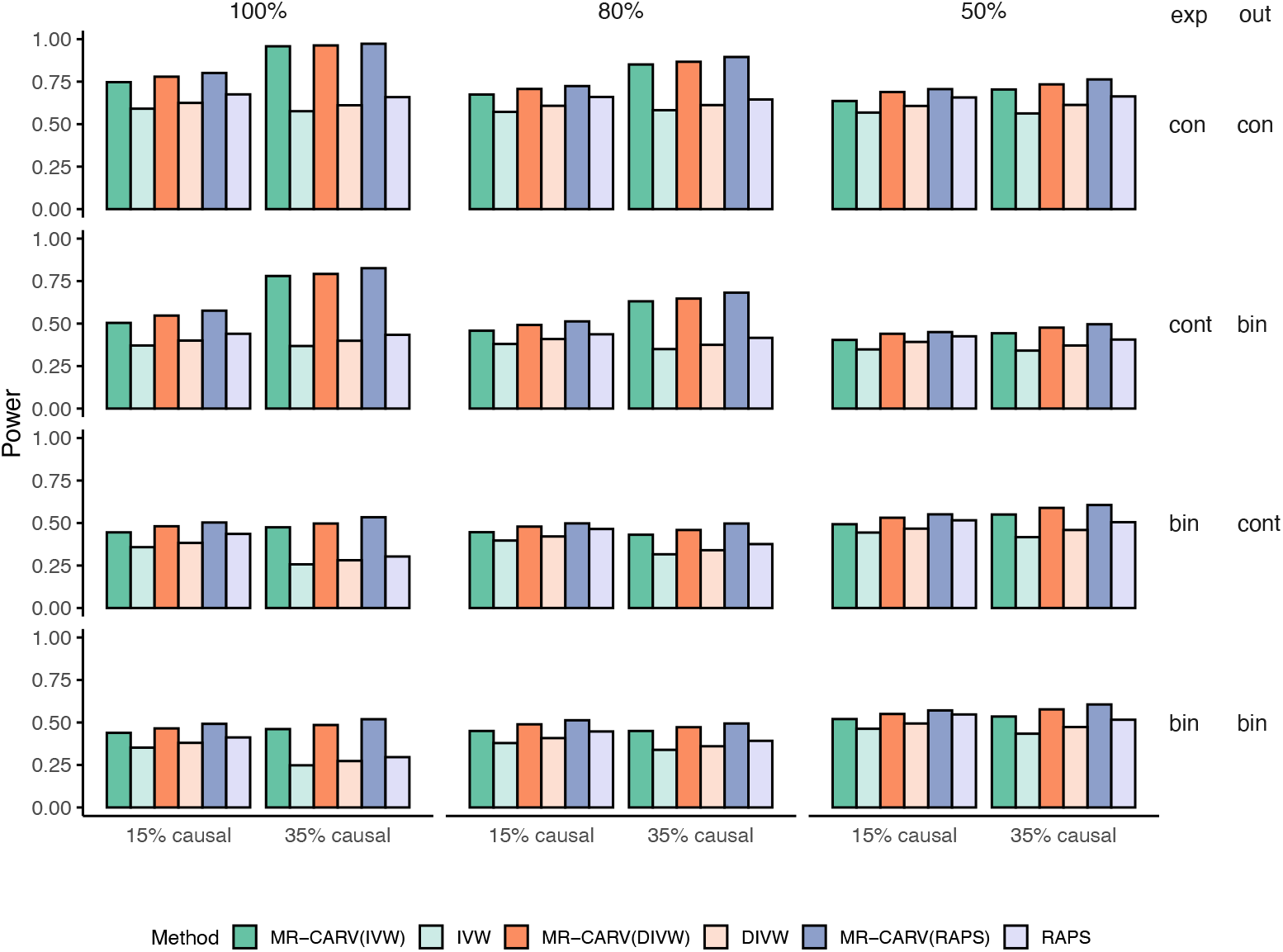
Power result for different methods under various exposure and outcome settings, based on 1000 simulations with sample size 10,000 for both exposure and outcome. The first row displays results for continuous exposure and continuous outcome with *β* = 0.04. The second row shows continuous exposure and binary outcome with *β* = 0.06. The third row presents binary exposure and continuous outcome with *β* = 0.2. The fourth row illustrates binary exposure and binary outcome with *β* = 0.4. Each column represents scenarios where 100%, 80%, and 50% of the variants have positive effects on the exposure. The X-axis shows the proportions of causal variants in the signal region, with 15% and 35% causal variants, respectively. The last two columns display the data types for exposure (exp) and outcome (out), respectively (cont: continuous, bin: binary). ALT Text: Bar plots displaying statistical power across MR methods under various simulation conditions with non-zero causal effect. MR-CARV shows consistently higher power.

For binary exposure and continuous outcome (*β* = 0.2), the MR-CARV methods (MRCARV(IVW), MR-CARV(dIVW), and MR-CARV(RAPS)) consistently exhibited higher statistical power compared to the existing methods (IVW, dIVW, and RAPS) (the third row in **Figure 2**). And the difference in statistical power between MR-CARV methods and existing methods still increased as causal variant ratio and positive effect ratio increase. However, unlike the previous settings, existing methods demonstrated higher statistical power with smaller causal and positive ratios of rare variants. For MR-CARV methods, the statistical power slightly increased with higher causal ratios, except when the positive ratio is 80%. This is likely due to the increased variability and noise introduced by higher causal and positive ratios of rare variants when simulating *X* and *Y*, although existing methods only consider common variants in the analysis. The binary nature of the exposure adds additional complexity because of the logit transformation, which may lead to issues such as non-collapsibility. The existing methods, which are not optimized to handle this increased noise, may struggle to detect true causal effects, resulting in lower statistical power. While MR-CARV methods utilize rare variant information, the non-collapsibility issues still presents challenges that can affect the estimates. However, including rare variants improves performance compared to existing methods. In the case of binary exposure and binary outcome (*β* = 0.4), it demonstrates similar performance as in the binary exposure and continuous outcome setting (the fourth row in **Figure 2**).

The power performance under negative true effect size (Figure S2) is similar to those in **Figure 2**. Figures S3 S14 demonstrate that MR-CARV methods still maintained correct type I error rate and had higher statistical power compared to existing methods even under small sample size. Although a higher sample size generally leads to higher statistical power, increasing the sample size of the outcome notably enhances statistical power, while increasing the sample size of the exposure has a less impact on statistical power. The details of MR-CARV evaluation under LD structure are provided in Supplementary Materials Section 5. MR-CARV consistently maintained well-controlled type I error and demonstrated improved power compared to existing MR methods that use only common variants. Similarly, the evaluation of MR-CARV under horizontal pleiotropy is presented in Supplementary Materials Section 6. MR-CARV maintained appropriate type I error under InSIDE-valid and balanced pleiotropy scenarios, and achieved higher statistical power than existing methods, consistent with the respective assumptions of each method.

### 3.2 Application to real data

To demonstrate the practical utility of our proposed MR-CARV framework, we applied the existing methods (IVW, dIVW, and RAPS), and MR-CARV methods (MR-CARV(IVW), MRCARV(dIVW), and MR-CARV(RAPS)), to explore the causal effects of lipids on preeclampsia. We utilized summary statistics for lipid traits, including High-Density Lipoprotein Cholesterol (HDL-C), Low-Density Lipoprotein Cholesterol (LDL-C), Triglycerides (TG), and Total Cholesterol (TC), from Selvaraj et al. [31]. The STAARpipeline was applied to perform both common and rare variant analyses, investigating their effects on blood lipid levels in a cohort of over 66,000 individuals in their paper. For rare variants, we included gene-coding, gene-noncoding, and non-gene regions. Here, we only set (*a*_1_, *a*_2_) = (1, 1) and do not include functional annotations in the weighting scheme, because Selvaraj et al. [31] presented only the summary effect size of rare variants with Burden(1,1). The preeclampsia data used in our analysis were obtained from the Nulliparous Pregnancy Outcomes Study: Monitoring Mothers-to-Be Heart Health Study (nuMoM2b-HHS) [32]. The nuMoM2b-HHS dataset provided individual-level whole genome sequencing and phenotype data. We used the STAARpipeline to assess the association between preeclampsia and both common and rare variants (i.e.MAF *<* 0.05), on 486 preeclampsia cases and 2,821 controls. Covariates included maternal age, age squared, the first 10 principal components calculated by genotypic data, race, and study sites [33]. A genetic relationship matrix was included as a random effect. We only selected uncorrelated common variants and Burden variables from rare variant sets as instruments.

Our analysis revealed a consistent negative association between HDL-C levels and preeclampsia, suggesting that higher HDL-C may confer a protective effect against the development of this condition (**Table 1**). When incorporating rare variants using the MR-CARV framework, the effect estimate remained similar but had a smaller standard error and narrower 95% confidence interval, reflecting increased precision. This improvement can be attributed to the additional genetic information contributed by rare variants, which enhances statistical power. Notably, the P-value of IVW decreased from 0.0659 (common variants only) to 0.0470 (common + rare variants), further supporting the benefit of including rare variants in MR analysis.

**Table 1.** Estimates, standard errors (SE), 95% confidence interval (CI), and P-values for the causal effect of HDL-C on preeclampsia from different methods.

| Method | Estimate | SE | 95% CI | P-value |
| --- | --- | --- | --- | --- |
| MR-CARV(IVW) | -0.020 | 0.0102 | [-0.0401, -0.0003] | 0.0470 |
| IVW | -0.023 | 0.0123 | [-0.0469, 0.0015] | 0.0659 |
| MR-CARV(dIVW) | -0.021 | 0.0104 | [-0.0408, -0.0003] | 0.0472 |
| dIVW | -0.023 | 0.0125 | [-0.0476, 0.0015] | 0.0660 |
| MR-CARV(RAPS) | -0.020 | 0.0103 | [-0.0406, -0.0002] | 0.0472 |
| RAPS | -0.023 | 0.0125 | [-0.0475, 0.0015] | 0.0660 |

**Figure 3** displays the estimates for IVW and MR-CARV(IVW), which appear similar. However, the higher precision of MR-CARV(IVW) makes the estimate more statistically significant. With the graph-based method to select the uncorrelated instruments, we identified 9 instruments in gene-coding regions, 1 instruments in gene-noncoding regions, 2 instruments in non-gene regions, and 63 instruments from common variants. Including these 12 instruments from rare variants already increased the statistical power of our analysis, and we could expect that incorporating more rare variant instruments will further enhance the statistical power. Notably, the summary statistics for rare variants are more dispersed compared to those for common variants, particularly for HDL-C. This greater spread is likely due to the smaller number of rare variants or the larger variance associated with the Burden variable in the set. The effect estimates of LDL-C, TG, and TC on the preeclampsia are shown in Table S2 in the Supplementary Materials. All the P values are higher than 0.05 either using the MR-CARV methods or the existing methods.

**Figure 3.**
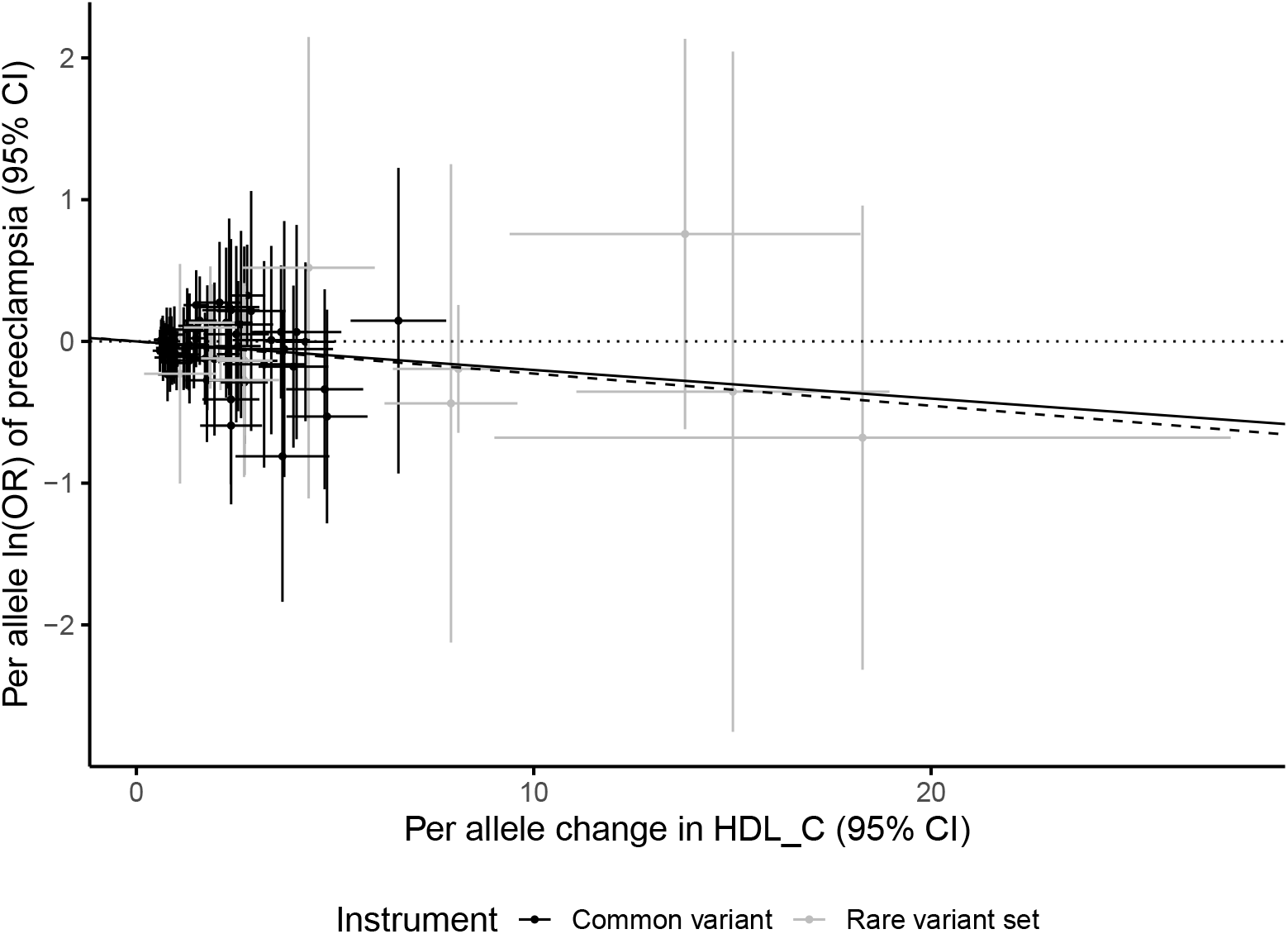
MR scatter plot illustrating the relationship between HDL-C and preeclampsia. The X-axis represents the per allele change in HDL-C (95% CI) from the GWAS, while the Y-axis represents the per allele ln(OR) for preeclampsia (95% CI) from the GWAS. Common variant statistics are depicted in black, and rare variant set statistics are shown in grey. The solid black line indicates the MR-CARV(IVW) estimate, and the dashed line represents the IVW estimate. Both the solid and dashed lines pass through point (0,0). ALT Text: Scatter plot of per allele HDL-C effects versus log-odds for preeclampsia. Includes confidence intervals and fitted lines from IVW and MR-CARV(IVW), comparing causal effect estimates using common vs. rare variants.

## 4 Discussion

In this study, we developed a novel two-sample MR framework, MR-CARV, which incorporates annotation-weighted rare variants in addition to the common variants, enhancing the performance of existing two-sample MR methods. Rare variants were aggregated into Burden variables using functional annotations and MAF-based weighting, similar to the STAAR approach [21], and the Cauchy combination test was applied to integrate results across multiple annotation-informed models. Extensive simulations demonstrated that MR-CARV framework effectively controls type I error and achieves higher statistical power than existing methods, particularly in settings with binary exposures and limited instrument strength from common variants. We also found that increasing the outcome sample size yields greater power gains than increasing the exposure sample size.

Applying the MR-CARV framework, we found that HDL-C shows a statistically significant protective effect against preeclampsia only when both common and rare variants are included. Incorporating rare variants led to more precise estimates, as indicated by smaller standard errors, even though fewer rare variant sets were used compared to common variants. This suggests potential for further gains with more rare variant data. Statistically, the estimated effect was marginally significant but consistent with prior studies [19, 34, 35]. HDL-C’s known anti-inflammatory and antioxidant properties [36, 37] may reduce oxidative stress and improve endothelial function, which are critical in preventing preeclampsia [38].

The growing availability of WGS data from large-scale initiatives such as the UK Biobank [28], All of Us [39], TOPMed [40], and the Million Veteran Program [41] has enabled broader analysis of rare variants. This expansion is supported by advanced tools like STAARpipeline [15], SAIGE [42, 43], and REGENIE [44]. MR-CARV currently leverages STAARpipeline to construct annotation-informed burden scores for rare variant sets. As WGS-based summary statistics become increasingly available, MR-CARV is well positioned to capitalize on this data. Moreover, our framework is flexible and can incorporate outputs from other rare variant association tools such as REGENIE.

Similar to existing MR studies [45, 46], the MR-CARV framework relies on the same key assumptions for valid causal inference. For example, to ensure the independence of instruments, we use an automated graph-based method to calculate correlations and only select uncorrelated instruments. However, more advanced techniques, such as Bayesian approaches, may offer better solutions in complex scenarios [47]. To address horizontal pleiotropy, the MR-CARV framework can be extended to incorporate rare variants into existing methods designed to account for pleiotropy, such as MR-Egger regression [48] and multivariable MR [49]. Overall, MR-CARV is a highly flexible tool that integrates both common and rare variants, offering a unified framework for causal inference in observational studies. Its adaptability allows the incorporation of advanced methodologies to address complex challenges, making it a valuable resource for exploring causal relationships between risk factors and complex diseases.

## 5 Key points

- The MR-CARV framework addresses key limitations in Mendelian Randomization (MR) by integrating both common and rare variants into established methods like IVW, dIVW, and RAPS, enhancing their statistical power.
- Rare variants are incorporated into the MR-CARV framework by weighting them with functional annotations, improving the biological relevance and precision of the analysis.
- The MR-CARV framework is highly flexible and can be adapted to extend other twosample MR methods, providing broad applicability across traits and diseases.
- We offer a publicly available R package, mr.carv, at https://github.com/yu-zhang-oYo/mr.carv which enables researchers to integrate rare and common variants seamlessly into MR analyses.
- Applying MR-CARV to data on blood lipid levels and preeclampsia reveals a significant protective effect of HDL-C on preeclampsia risk, demonstrating its practical utility in uncovering meaningful causal relationships.

## Data Availability

Access to the nuMoM2b-HHS individual-level whole genome sequencing and phenotype data is
available through dbGaP at dbGaP study ID phs002808.v1.p1

https://github.com/yu-zhang-oYo/mr.carv

## 6 Supplementary information

Supplementary information can be found at file supplements.

## 7 Data availability

The summary statistics for lipid traits are available at the published paper Selvaraj et al. [31]. Access to the nuMoM2b-HHS individual-level whole genome sequencing and phenotype data is available through dbGaP at dbGaP study ID phs002808.v1.p1.

## 8 Code availability

MR-CARV is implemented as an open source R package available at https://github.com/yu-zhang-oYo/mr.carv.

## 10 Acknowledgements

The authors would like to thank Stephen Burgess, PhD and Xiaofeng Zhu, PhD for their insightful discussions. This research was supported in part by Lilly Endowment, Inc., through its support for the Indiana University Pervasive Technology Institute.

## 11 Author Contributions

Conceptualization, Q.Y. and Y.Z.; Methodology, Q.Y. and Y.Z.; Investigation, Y.Z., Q.Y. and X.L.; Software, Y.Z., Writing – Original Draft, Y.Z; Writing – Review & Editing, Y.Z., Q.Y., X.L., N.L., M.L., D.H., C.B., T.W., K.R., J.C., L.L., A.F., G.S.; Supervision, Q.Y. and N.L.

## 13 Declaration of generative AI and AI-assisted technologies in the writing process

During the preparation of this work the authors used ChatGPT (OpenAI, San Francisco, CA) in order to refine language and improve the clarity of the manuscript. After using this tool, the authors reviewed and edited the content as needed and take full responsibility for the content of the publication.

## References

[1] Xu, J., Li, M., Gao, Y., et al. Using Mendelian randomization as the cornerstone for causal inference in epidemiology. In: Environmental Science and Pollution Research 29.4 (2022), pp. 5827–5839. doi: 10.1007/s11356-021-15939-3.

[2] Sanderson, E., Glymour, M. M., Holmes, M. V., et al. Mendelian randomization. In: Nature Reviews Methods Primers 2.1 (2022), p. 6. doi: 10.1038/s43586-021-00092-5.

[3] Lee, S., Abecasis, G. R., Boehnke, M., et al. Rare-variant association analysis: study designs and statistical tests. In: The American Journal of Human Genetics 95.1 (2014), pp. 5–23. doi: 10.1016/j.ajhg.2014.06.009.

[4] Wang, Q., Dhindsa, R. S., Carss, K., et al. Rare variant contribution to human disease in 281,104 UK Biobank exomes. In: Nature 597.7877 (2021), pp. 527–532. doi: 10.1038/s41586-021-03855-y.

[5] Ye, T., Shao, J., and Kang, H. Debiased inverse-variance weighted estimator in twosample summary-data Mendelian randomization. In: The Annals of statistics 49.4 (2021), pp. 2079–2100. doi: 10.1214/20-AOS2027.

[6] Madsen, B. E. and Browning, S.R. A groupwise association test for rare mutations using a weighted sum statistic. In: PLoS genetics 5.2 (2009), e1000384. doi: 10.1371/journal.pgen.1000384.

[7] Wu, M. C., Lee, S., Cai, T., et al. Rare-variant association testing for sequencing data with the sequence kernel association test. In: The American Journal of Human Genetics 89.1 (2011), pp. 82–93. doi: 10.1016/j.ajhg.2011.05.029.

[8] Liu, Y., Chen, S., Li, Z., et al. ACAT: a fast and powerful p value combination method for rare-variant analysis in sequencing studies. In: The American Journal of Human Genetics 104.3 (2019), pp. 410–421. doi: 10.1016/j.ajhg.2019.01.002.

[9] Tada, H., Kawashiri, M.-a., and Yamagishi, M. Comprehensive genotyping in dyslipidemia: mendelian dyslipidemias caused by rare variants and Mendelian randomization studies using common variants. In: Journal of human genetics 62.4 (2017), pp. 453–458. doi: 10.1038/jhg.2016.159.

[10] Rao, A. R. and Nelson, S. F. Calculating the statistical significance of rare variants causal for Mendelian and complex disorders. In: BMC medical genomics 11 (2018), pp. 1–17. doi: 10.1186/s12920-018-0371-9.

[11] Hsu, L.-A., Teng, M.-S., Wu, S., et al. Common and rare PCSK9 variants associated with low-density lipoprotein cholesterol levels and the risk of diabetes mellitus: A mendelian randomization study. In: International Journal of Molecular Sciences 23.18 (2022), p. 10418. doi: 10.3390/ijms231810418.

[12] Triozzi, J. L., Hsi, R. S., Wang, G., et al. Mendelian randomization analysis of genetic proxies of thiazide diuretics and the reduction of kidney stone risk. In: JAMA network open 6.11 (2023), e2343290–e2343290. doi: 10.1001/jamanetworkopen.2023.43290.

[13] Abdi, G., Tarighat, M.A., Jain, M., et al. “Revolutionizing Genomics: Exploring the Potential of Next-Generation Sequencing”. In: Advances in Bioinformatics. Springer, 2024, pp. 1–33. doi: 10.1007/978-981-99-8401-5_1.

[14] Austin, M. A., Hair, M. S., and Fullerton, S. M. Research guidelines in the era of large-scale collaborations: an analysis of Genome-wide Association Study Consortia. In: American journal of epidemiology 175.9 (2012), pp. 962–969. doi: 10.1093/aje/kwr441.

[15] Li, Z., Li, X., Zhou, H., et al. A framework for detecting noncoding rare-variant associations of large-scale whole-genome sequencing studies. In: Nature methods 19.12 (2022), pp. 1599–1611. doi: 10.1038/s41592-022-01640-x.

[16] Yang, Y., Wang, Y., Lv, Y., et al. Dissecting the roles of lipids in preeclampsia. In: Metabolites 12.7 (2022), p. 590. doi: 10.3390/metabo12070590.

[17] Zhou, X., Han, T.-L., Chen, H., et al. Impaired mitochondrial fusion, autophagy, biogenesis and dysregulated lipid metabolism is associated with preeclampsia. In: Experimental cell research 359.1 (2017), pp. 195–204. doi: 10.1016/j.yexcr.2017.07.029.

[18] Spracklen, C. N., Smith, C. J., Saftlas, A. F., et al. Maternal hyperlipidemia and the risk of preeclampsia: a meta-analysis. In: American journal of epidemiology 180.4 (2014), pp. 346–358. doi: 10.1093/aje/kwu145.

[19] Spracklen, C. N., Saftlas, A. F., Triche, E. W., et al. Genetic predisposition to dyslipidemia and risk of preeclampsia. In: American journal of hypertension 28.7 (2015), pp. 915–923. doi: 10.1093/ajh/hpu242.

[20] Burgess, S., Butterworth, A., and Thompson, S. G. Mendelian randomization analysis with multiple genetic variants using summarized data. In: Genetic epidemiology 37.7 (2013), pp. 658–665. doi: 10.1002/gepi.21758.

[21] Li, X., Li, Z., Zhou, H., et al. Dynamic incorporation of multiple in silico functional annotations empowers rare variant association analysis of large whole-genome sequencing studies at scale. In: Nature genetics 52.9 (2020), pp. 969–983. doi: 10.1038/s41588-020-0676-4.

[22] Liu, Y. and Xie, J. Cauchy combination test: a powerful test with analytic p-value calculation under arbitrary dependency structures. In: Journal of the American Statistical Association (2020). doi: 10.1080/01621459.2018.1554485.

[23] Zhao, Q., Wang, J., Hemani, G., et al. Statistical inference in two-sample summary-data Mendelian randomization using robust adjusted profile score. In: The Annals of Statistics 48.3 (2020), pp. 1742–1769. doi: 10.1214/19-AOS1866.

[24] Burgess, S. and Thompson, S. G. Mendelian randomization: methods for causal inference using genetic variants. CRC Press, 2021.

[25] Swerdlow, D. I., Preiss, D., Kuchenbaecker, K. B., et al. HMG-coenzyme A reductase inhibition, type 2 diabetes, and bodyweight: evidence from genetic analysis and randomised trials. In: The Lancet 385.9965 (2015), pp. 351–361. doi: 10.1016/S0140-6736(14)61183-1.

[26] Schmidt, A. F., Finan, C., Gordillo-Marañón, M., et al. Genetic drug target validation using Mendelian randomisation. In: Nature communications 11.1 (2020), p. 3255. doi: 10.1038/s41467-020-16969-0.

[27] Tsukiyama, S., Ide, M., Ariyoshi, H., et al. A new algorithm for generating all the maximal independent sets. In: SIAM Journal on Computing 6.3 (1977), pp. 505–517. doi: 10.1137/020603.

[28] Hofmeister, R. J., Ribeiro, D. M., Rubinacci, S., et al. Accurate rare variant phasing of whole-genome and whole-exome sequencing data in the UK Biobank. In: Nature Genetics 55.7 (2023), pp. 1243–1249. doi: 10.1038/s41588-023-01415-w.

[29] Clarke, L., Zheng-Bradley, X., Smith, R., et al. The 1000 Genomes Project: data management and community access. In: Nature methods 9.5 (2012), pp. 459–462. doi: 10.1038/nmeth.1974.

[30] Dimitromanolakis, A., Xu, J., Krol, A., et al. sim1000G: a user-friendly genetic variant simulator in R for unrelated individuals and family-based designs. In: BMC bioinformatics 20 (2019), pp. 1–9. doi: 10.1186/s12859-019-2611-1.

[31] Selvaraj, M. S., Li, X., Li, Z., et al. Whole genome sequence analysis of blood lipid levels in 66,000 individuals. In: Nature communications 13.1 (2022), p. 5995. doi: 10.1038/s41467-022-33510-7.

[32] Haas, D. M., Ehrenthal, D. B., Koch, M. A., et al. Pregnancy as a window to future cardiovascular health: design and implementation of the nuMoM2b Heart Health Study. In: American journal of epidemiology 183.6 (2016), pp. 519–530. doi: 10.1093/aje/kwv309.

[33] Yan, Q., Blue, N. R., Truong, B., et al. Genetic Associations with Placental Proteins in Maternal Serum Identify Biomarkers for Hypertension in Pregnancy. In: medRxiv (2023), pp. 2023–05. doi: 10.1101/2023.05.25.23290460.

[34] Ding, Y., Yao, M., Liu, J., et al. Association between human blood metabolome and the risk of pre-eclampsia. In: Hypertension Research 47.4 (2024), pp. 1063–1072. doi: 10.1038/s41440-024-01586-x.

[35] Hosier, H., Lipkind, H. S., Rasheed, H., et al. Dyslipidemia and risk of preeclampsia: a multiancestry mendelian randomization study. In: Hypertension 80.5 (2023), pp. 1067– 1076. doi: 10.1161/HYPERTENSIONAHA.122.20426.

[36] Barter, P. J., Nicholls, S., Rye, K.-A., et al. Antiinflammatory properties of HDL. In: Circulation research 95.8 (2004), pp. 764–772. doi: 10.1161/01.RES.0000146094.59640.13.

[37] Tran-Dinh, A., Diallo, D., Delbosc, S., et al. HDL and endothelial protection. In: British journal of pharmacology 169.3 (2013), pp. 493–511. doi: 10.1111/bph.12174.

[38] Sánchez-Aranguren, L. C., Prada, C. E., Riaño-Medina, C. E., et al. Endothelial dysfunction and preeclampsia: role of oxidative stress. In: Frontiers in physiology 5 (2014), p. 372. doi: 10.3389/fphys.2014.00372.

[39] Us Research Program Investigators, A. of. The “All of Us” research program. In: New England Journal of Medicine 381.7 (2019), pp. 668–676. doi: 10.7759/cureus.65831.

[40] Taliun, D., Harris, D. N., Kessler, M. D., et al. Sequencing of 53,831 diverse genomes from the NHLBI TOPMed Program. In: Nature 590.7845 (2021), pp. 290–299. doi: 10.1038/s41586-021-03205-y.

[41] Gaziano, J. M., Concato, J., Brophy, M., et al. Million Veteran Program: A mega-biobank to study genetic influences on health and disease. In: Journal of clinical epidemiology 70 (2016), pp. 214–223. doi: 10.1016/j.jclinepi.2015.09.016.

[42] Zhou, W., Nielsen, J. B., Fritsche, L. G., et al. Efficiently controlling for case-control imbalance and sample relatedness in large-scale genetic association studies. In: Nature genetics 50.9 (2018), pp. 1335–1341. doi: 10.1038/s41588-018-0184-y.

[43] Zhou, W., Zhao, Z., Nielsen, J. B., et al. Scalable generalized linear mixed model for region-based association tests in large biobanks and cohorts. In: Nature genetics 52.6 (2020), pp. 634–639. doi: 10.1038/s41588-020-0621-6.

[44] Mbatchou, J., Barnard, L., Backman, J., et al. Computationally efficient whole-genome regression for quantitative and binary traits. In: Nature genetics 53.7 (2021), pp. 1097– 1103. doi: 10.1038/s41588-021-00870-7.

[45] VanderWeele, T. J., Tchetgen, E. J. T., Cornelis, M., et al. Methodological challenges in mendelian randomization. In: Epidemiology 25.3 (2014), pp. 427–435. doi: 10.1097/EDE.0000000000000081.

[46] Leeuw, C. de, Savage, J., Bucur, I. G., et al. Understanding the assumptions underlying Mendelian randomization. In: European Journal of Human Genetics 30.6 (2022), pp. 653– 660. doi: 10.1038/s41431-022-01038-5.

[47] Herce, Á. and Salvador, M. Instrument Selection in Panel Data Models with Endogeneity: A Bayesian Approach. In: Econometrics 12.4 (2024). issn: 2225-1146. doi: 10.3390/econometrics12040036.

[48] Burgess, S. and Thompson, S. G. Interpreting findings from Mendelian randomization using the MR-Egger method. In: European journal of epidemiology 32 (2017), pp. 377– 389. doi: 10.1007/s10654-017-0255-x.

[49] Rees, J. M., Wood, A. M., and Burgess, S. Extending the MR-Egger method for multivariable Mendelian randomization to correct for both measured and unmeasured pleiotropy. In: Statistics in medicine 36.29 (2017), pp. 4705–4718. doi: 10.1002/sim.7492.

