## supplement for "A novel two-sample Mendelian randomization framework integrating common and rare variants: application to assess the effect of HDL-C on preeclampsia risk" (from another version of this preprint: version 1, 2025-08-24)

### Supplementary Materials

#### 1 Supplementary Tables

**Table S1: Effect sizes ($\beta$) used for statistical power evaluation under different combinations of exposure and outcome types.**

| Exposure Type | Outcome Type | Sample Size: 10,000 | Sample Sizes: 1,000/2,000 |
|---|---|---|---|
| Continuous | Continuous | $\pm 0.04$ | $\pm 0.10$ |
| Continuous | Binary | $\pm 0.06$ | $\pm 0.20$ |
| Binary | Continuous | $\pm 0.20$ | $\pm 0.60$ |
| Binary | Binary | $\pm 0.40$ | $\pm 1.00$ |

**Table S2: Estimates, standard errors (SE), 95% confidence interval (CI) and P-values for the causal effect of HDL-C, LDL-C, TG, and TC on preeclampsia from different methods.**

| Exposure | Method | Estimate | SE | 95% CI | P-value |
|---|---|---|---|---|---|
| HDL-C | MR-CARV(IVW) | -0.0211 | 0.0101 | [-0.0410, -0.0013] | 0.0365 |
| | IVW | -0.0237 | 0.0123 | [-0.0478, 0.0004] | 0.0538 |
| | IVW (rare-only) | -0.0158 | 0.0178 | [-0.0507, 0.0191] | 0.3751 |
| | MR-CARV(dIVW) | -0.0217 | 0.0104 | [-0.0421, -0.0013] | 0.0368 |
| | dIVW | -0.0241 | 0.0125 | [-0.0485, 0.0004] | 0.0539 |
| | dIVW (rare-only) | -0.0167 | 0.0188 | [-0.0536, 0.0202] | 0.3761 |
| | MR-CARV(RAPS) | -0.0215 | 0.0105 | [-0.0420, -0.0010] | 0.0401 |
| | RAPS | -0.0240 | 0.0125 | [-0.0485, 0.0005] | 0.0548 |
| | RAPS (rare-only) | -0.0161 | 0.0191 | [-0.0536, 0.0213] | 0.3989 |
| LDL-C | MR-CARV(IVW) | 0.0014 | 0.0044 | [-0.0072, 0.0099] | 0.7489 |
| | IVW | -0.0013 | 0.0053 | [-0.0117, 0.0091] | 0.8033 |
| | IVW (rare-only) | 0.0104 | 0.0086 | [-0.0064, 0.0272] | 0.2243 |
| | MR-CARV(dIVW) | 0.0014 | 0.0044 | [-0.0072, 0.0099] | 0.7457 |
| | dIVW | -0.0013 | 0.0053 | [-0.0117, 0.0090] | 0.7998 |
| | dIVW (rare-only) | 0.0108 | 0.0089 | [-0.0067, 0.0282] | 0.2260 |
| | MR-CARV(RAPS) | 0.0014 | 0.0042 | [-0.0068, 0.0097] | 0.7349 |
| | RAPS | -0.0013 | 0.0048 | [-0.0107, 0.0080] | 0.7772 |
| | RAPS (rare-only) | 0.0106 | 0.0090 | [-0.0071, 0.0282] | 0.2394 |
| TG | MR-CARV(IVW) | 0.2179 | 0.3829 | [-0.5326, 0.9683] | 0.5694 |
| | IVW | 0.1964 | 0.4020 | [-0.5914, 0.9843] | 0.6251 |
| | IVW (rare-only) | 0.4455 | 1.3793 | [-2.2579, 3.1488] | 0.7467 |
| | MR-CARV(dIVW) | 0.2223 | 0.3870 | [-0.5362, 0.9808] | 0.5657 |
| | dIVW | 0.1994 | 0.4035 | [-0.5915, 0.9903] | 0.6213 |
| | dIVW (rare-only) | 0.4823 | 1.3753 | [-2.2133, 3.1779] | 0.7258 |
| | MR-CARV(RAPS) | 0.2216 | 0.3795 | [-0.5222, 0.9653] | 0.5593 |
| | RAPS | 0.1994 | 0.3944 | [-0.5736, 0.9723] | 0.6131 |
| | RAPS (rare-only) | 0.4557 | 1.3961 | [-2.2807, 3.1921] | 0.7441 |
| TC | MR-CARV(IVW) | 0.0042 | 0.0037 | [-0.0031, 0.0114] | 0.2581 |
| | IVW | 0.0020 | 0.0045 | [-0.0068, 0.0109] | 0.6508 |
| | IVW (rare-only) | 0.0099 | 0.0068 | [-0.0035, 0.0233] | 0.1488 |
| | MR-CARV(dIVW) | 0.0043 | 0.0039 | [-0.0033, 0.0119] | 0.2707 |
| | dIVW | 0.0021 | 0.0045 | [-0.0068, 0.0110] | 0.6479 |
| | dIVW (rare-only) | 0.0102 | 0.0072 | [-0.0038, 0.0243] | 0.1527 |
| | MR-CARV(RAPS) | 0.0043 | 0.0037 | [-0.0029, 0.0114] | 0.2447 |
| | RAPS | 0.0021 | 0.0043 | [-0.0063, 0.0104] | 0.6252 |
| | RAPS (rare-only) | 0.0100 | 0.0072 | [-0.0042, 0.0243] | 0.1659 |

*Note.* IVW, dIVW, and RAPS refer to existing MR methods using only common variants; rare-only versions use the same methods applied to rare variant burden variables.

#### 2 Power results with sample size 10,000 and negative true effect size

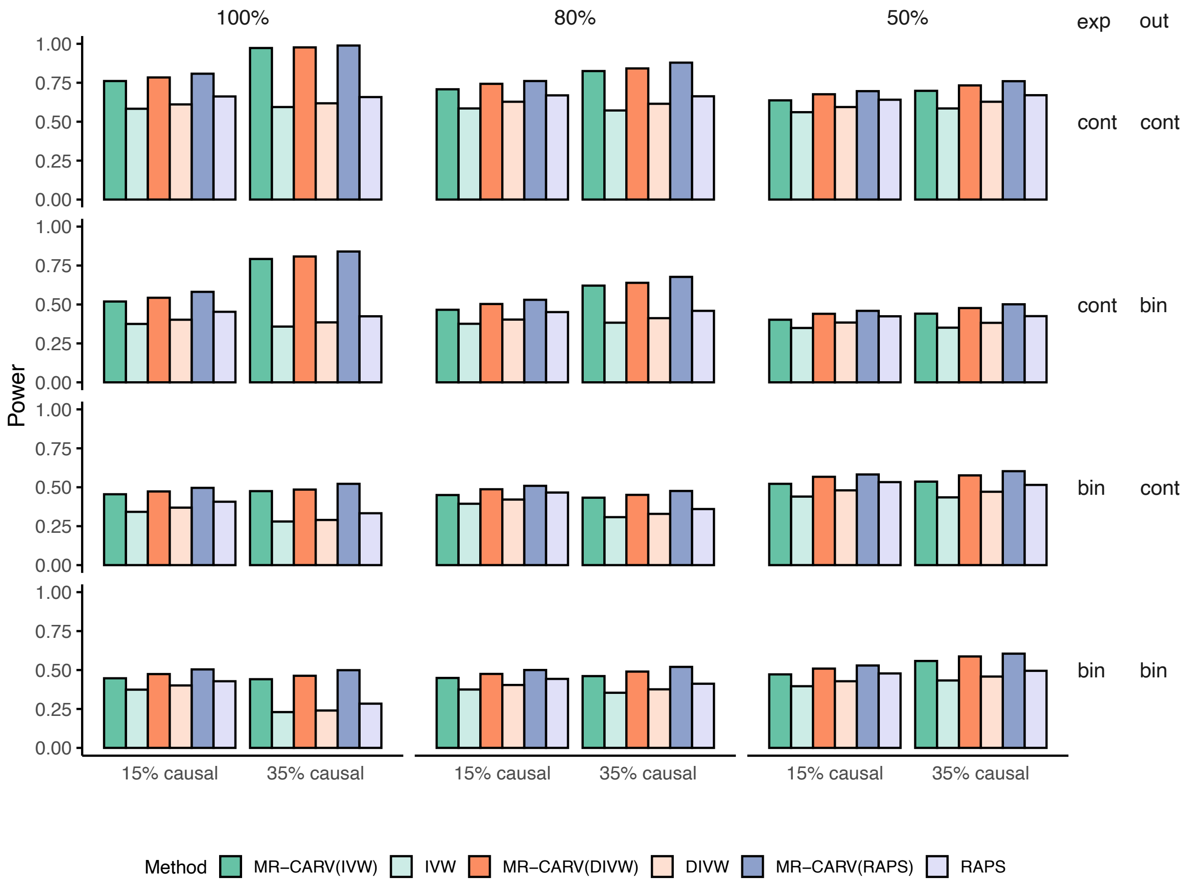


**Figure S1: Power results for different methods under various exposure and outcome settings, based on 1000 simulations with sample size 10,000 for both exposure and outcome.** The first row displays results for continuous exposure and continuous outcome with $\beta = -0.04$. The second row shows continuous exposure and binary outcome with $\beta = -0.06$. The third row presents binary exposure and continuous outcome with $\beta = -0.2$. The fourth row illustrates binary exposure and binary outcome with $\beta = -0.4$. Each column represents scenarios where 100%, 80%, and 50% of the variants have positive effects on the exposure. The X-axis shows the proportions of causal variants in the signal region, with 15% and 35% causal variants, respectively. The last two columns display the data types for exposure (exp) and outcome (out), respectively (cont: continuous, bin: binary).

### 3 simulation results under small sample size

#### 3.1 Continuous Exposure, Continuous Outcome

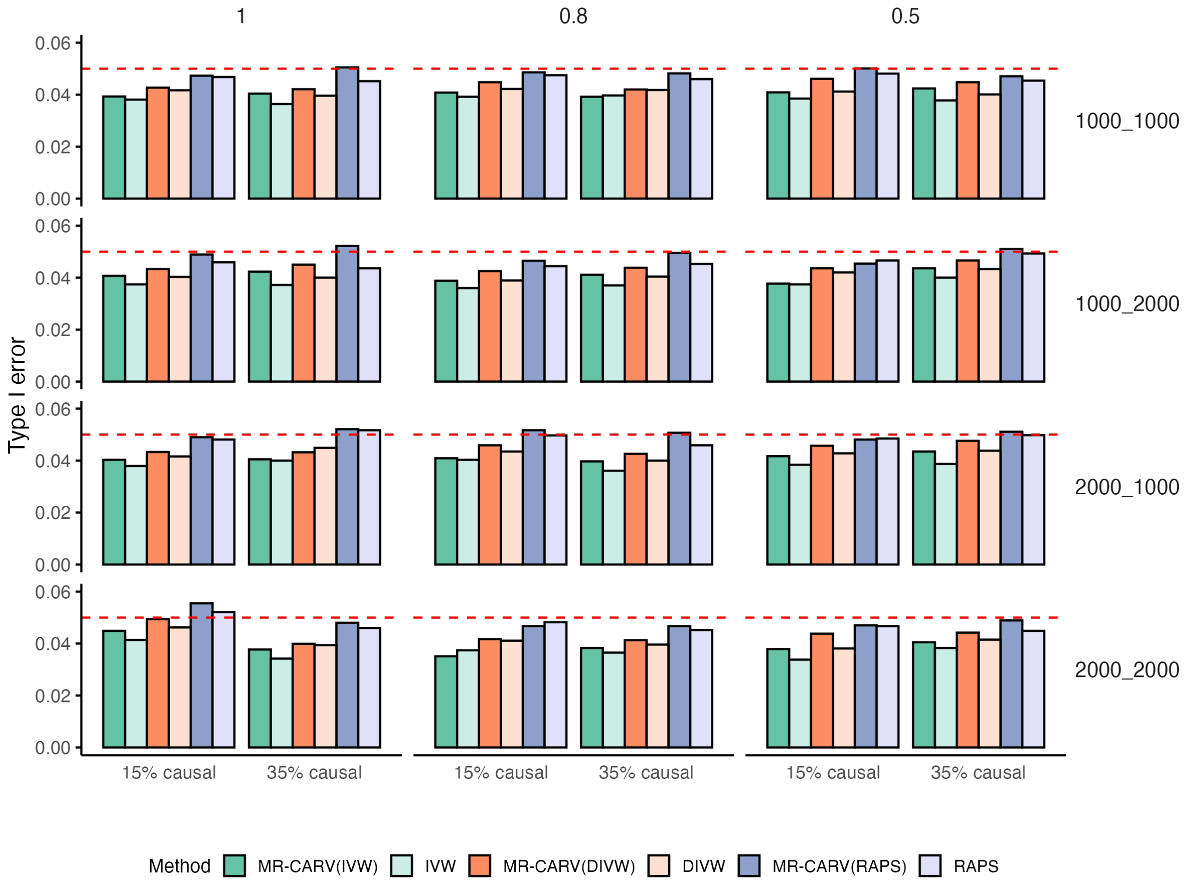


**Figure S2: Type I error rate under 10,000 simulations for continuous exposure and continuous outcome.** The first row represents sample size 1,000 for both exposure and outcome, the second row represents sample size 1,000 for exposure and sample size 2,000 for outcome, the third row represents sample size 2,000 for exposure and sample size 1,000 for outcome, and the fourth row represents sample size 2,000 for both exposure and outcome. The three columns represent the scenarios where 100%, 80%, and 50% of the variants have positive effects on the exposure. The X axis shows the proportions of causal variants in the signal region, with 15% and 35% causal variants respectively.

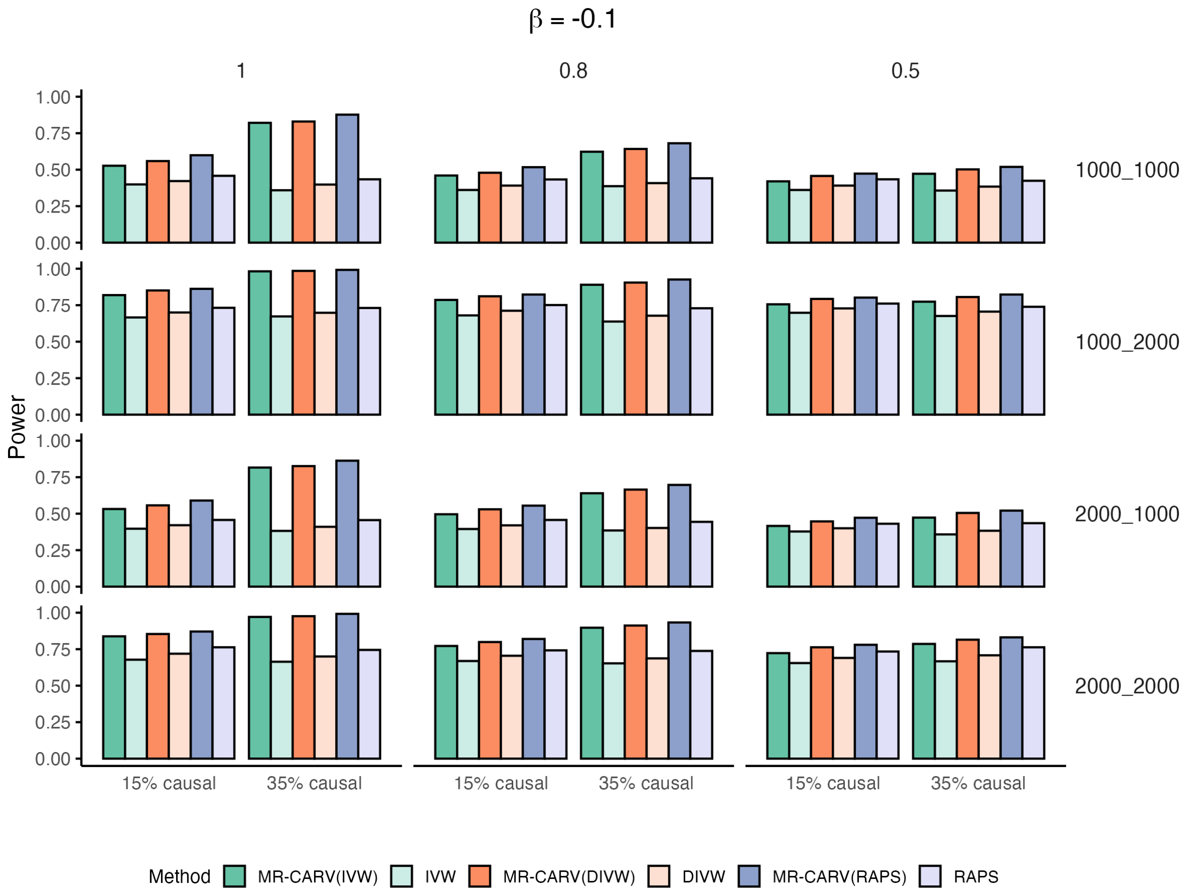


**Figure S3: Power results under 1,000 simulations when $\beta = -0.1$ for continuous exposure and continuous outcome.** The first row represents sample size 1,000 for both exposure and outcome, the second row represents sample size 1,000 for exposure and sample size 2,000 for outcome, the third row represents sample size 2,000 for exposure and sample size 1,000 for outcome, and the fourth row represents sample size 2,000 for both exposure and outcome. The three columns represent the scenarios where 100%, 80%, and 50% of the variants have positive effects on the exposure. The X axis shows the proportions of causal variants in the signal region, with 15% and 35% causal variants respectively.

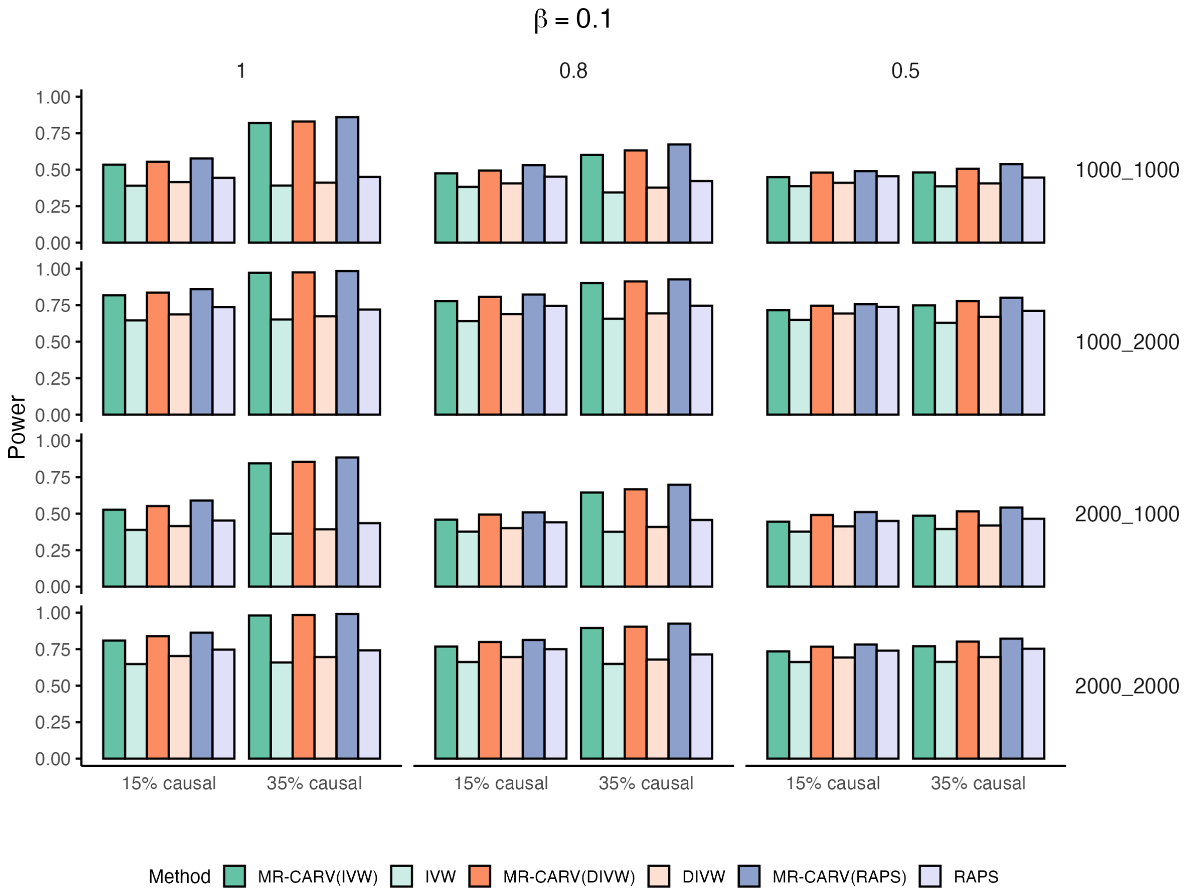


**Figure S4: Power results under 1,000 simulations when $\beta = 0.1$ for continuous exposure and continuous outcome.** The first row represents sample size 1,000 for both exposure and outcome, the second row represents sample size 1,000 for exposure and sample size 2,000 for outcome, the third row represents sample size 2,000 for exposure and sample size 1,000 for outcome, and the fourth row represents sample size 2,000 for both exposure and outcome. The three columns represent the scenarios where 100%, 80%, and 50% of the variants have positive effects on the exposure. The X axis shows the proportions of causal variants in the signal region, with 15% and 35% causal variants respectively.

#### 3.2 Continuous Exposure, Binary Outcome

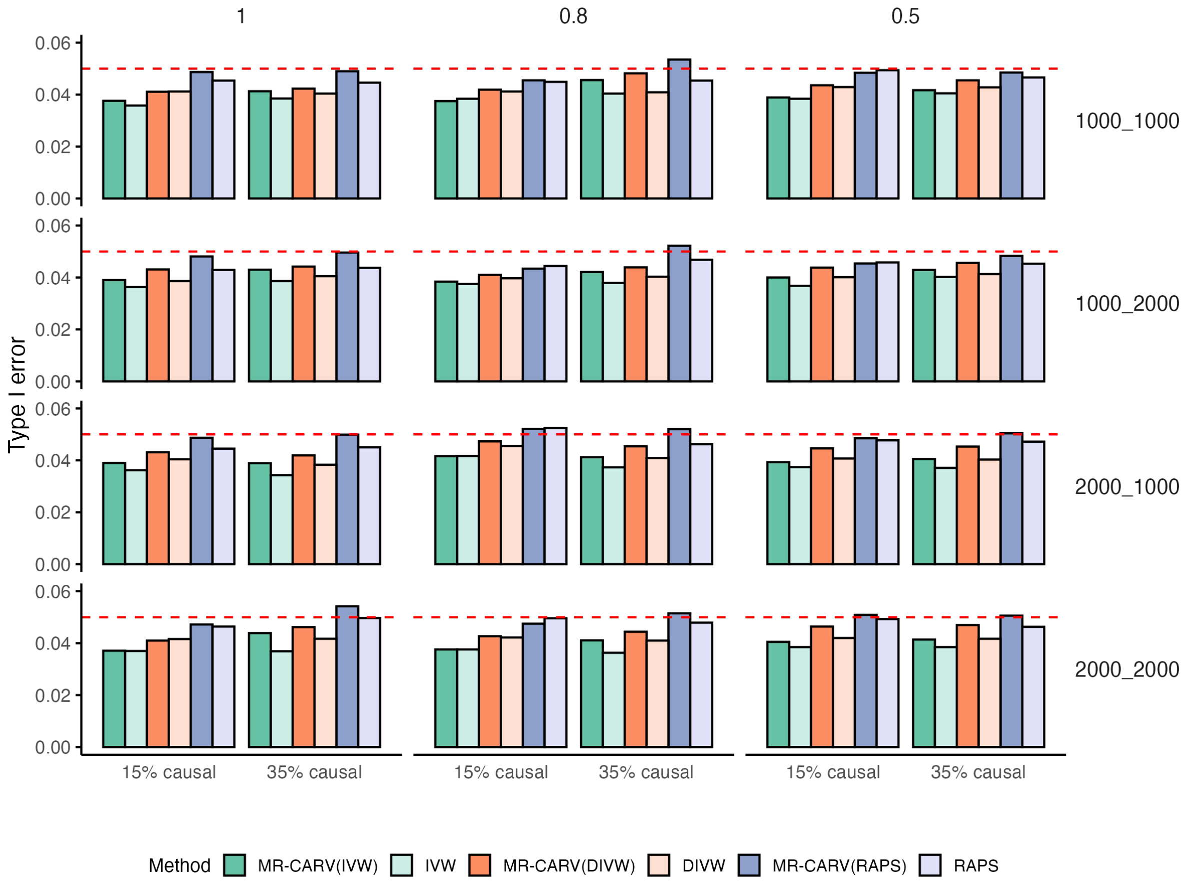


**Figure S5: Type I error rate under 10,000 simulations for continuous exposure and binary outcome.** The first row represents sample size 1,000 for both exposure and outcome, the second row represents sample size 1,000 for exposure and sample size 2,000 for outcome, the third row represents sample size 2,000 for exposure and sample size 1,000 for outcome, and the fourth row represents sample size 2,000 for both exposure and outcome. The three columns represent the scenarios where 100%, 80%, and 50% of the variants have positive effects on the exposure. The X axis shows the proportions of causal variants in the signal region, with 15% and 35% causal variants respectively.

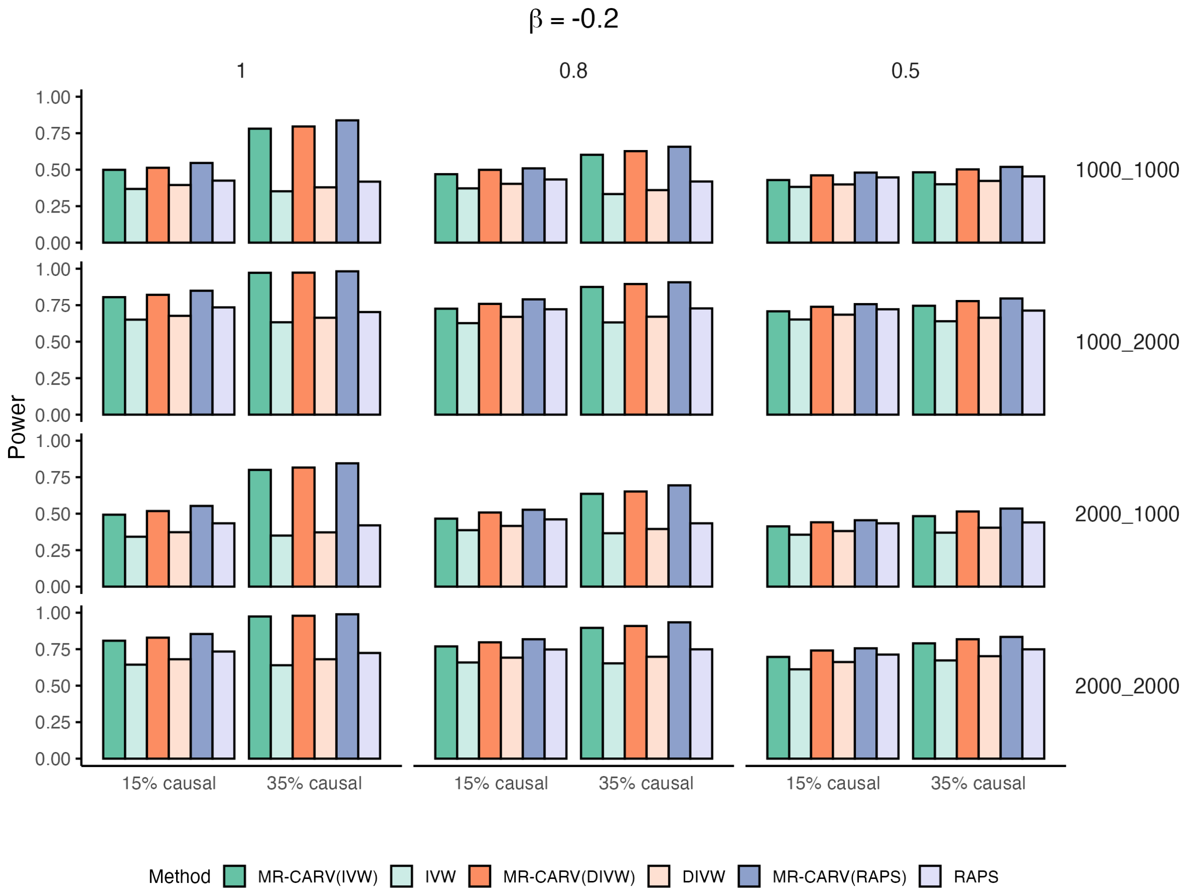


**Figure S6: Power results under 1,000 simulations when $\beta = -0.2$ for continuous exposure and binary outcome.** The first row represents sample size 1,000 for both exposure and outcome, the second row represents sample size 1,000 for exposure and sample size 2,000 for outcome, the third row represents sample size 2,000 for exposure and sample size 1,000 for outcome, and the fourth row represents sample size 2,000 for both exposure and outcome. The three columns represent the scenarios where 100%, 80%, and 50% of the variants have positive effects on the exposure. The X axis shows the proportions of causal variants in the signal region, with 15% and 35% causal variants respectively.

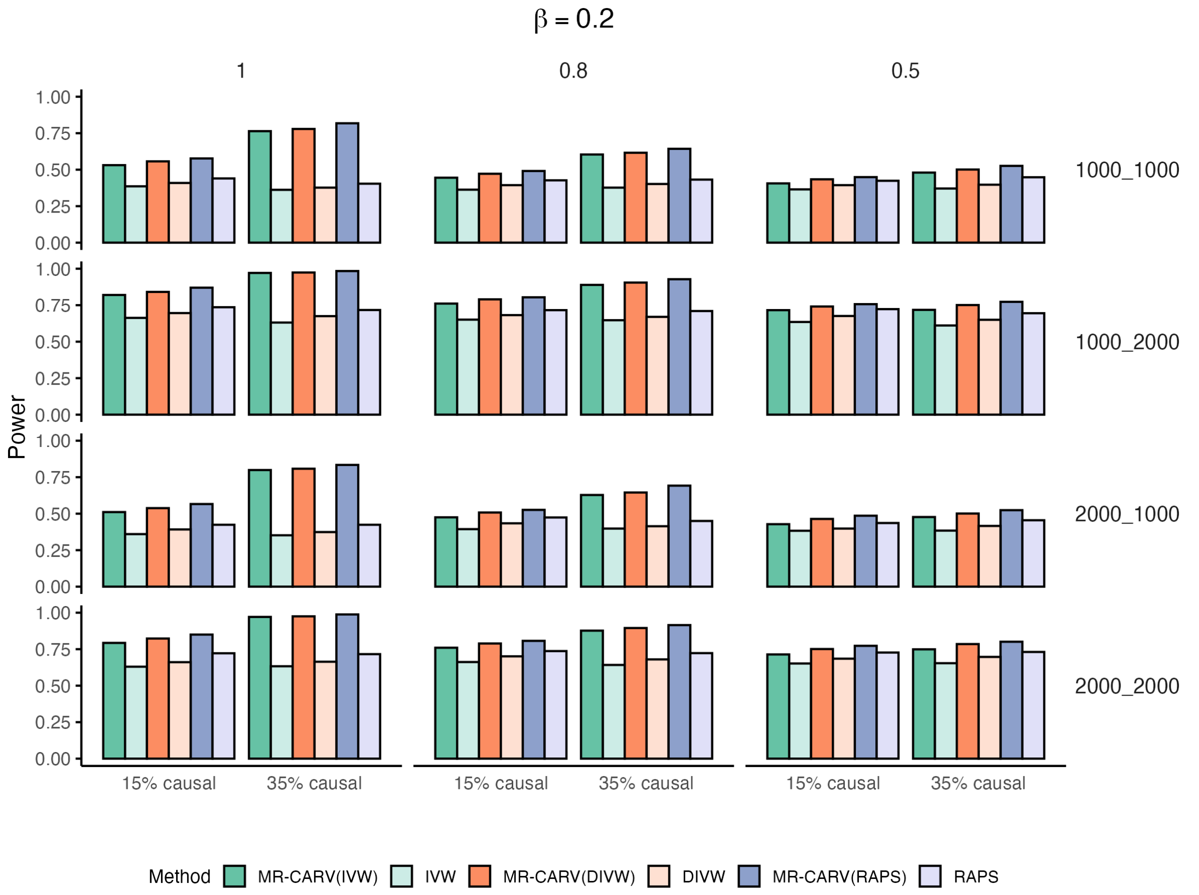


**Figure S7: Power results under 1,000 simulations when $\beta = 0.2$ for continuous exposure and binary outcome.** The first row represents sample size 1,000 for both exposure and outcome, the second row represents sample size 1,000 for exposure and sample size 2,000 for outcome, the third row represents sample size 2,000 for exposure and sample size 1,000 for outcome, and the fourth row represents sample size 2,000 for both exposure and outcome. The three columns represent the scenarios where 100%, 80%, and 50% of the variants have positive effects on the exposure. The X axis shows the proportions of causal variants in the signal region, with 15% and 35% causal variants respectively.

8 ### 3.3 Binary Exposure, Continuous Outcome

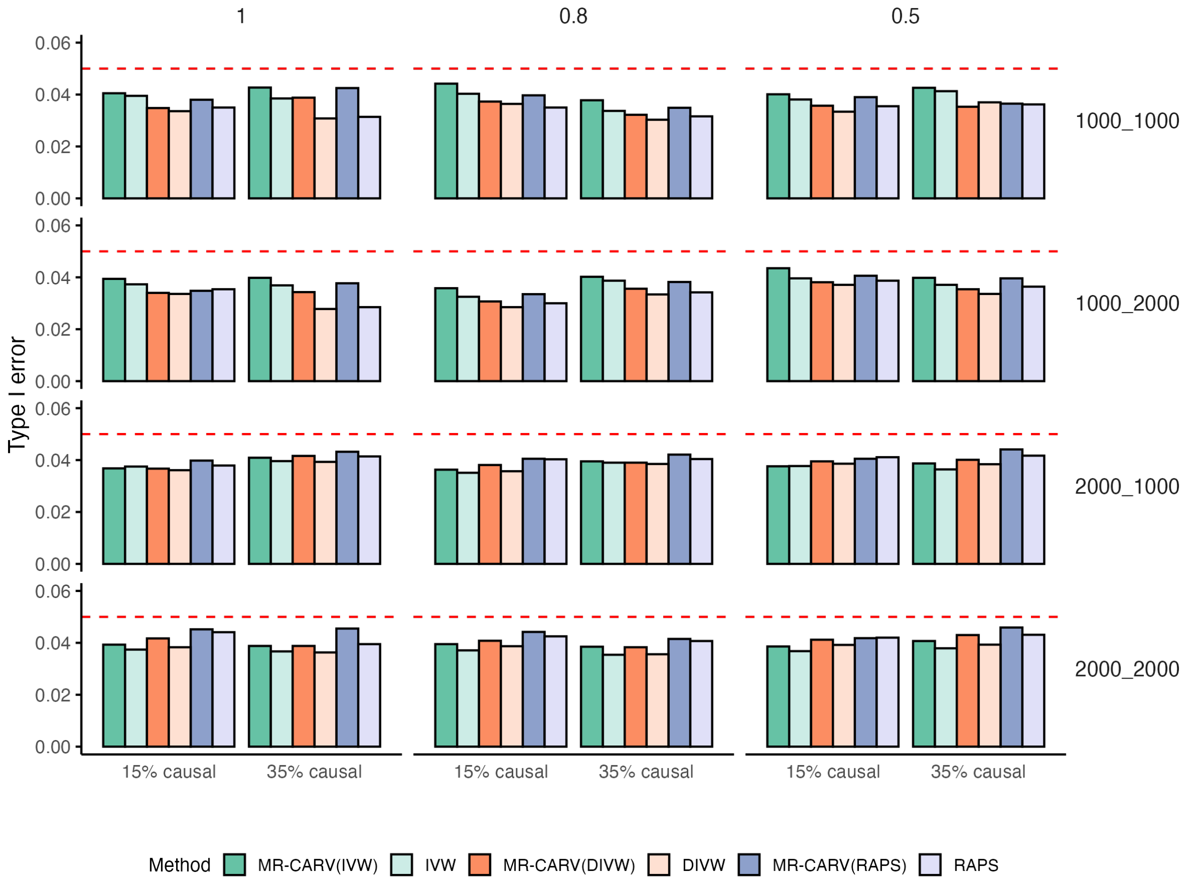


**Figure S8: Type I error rate under 10,000 simulations for binary exposure and continuous outcome.** The first row represents sample size 1,000 for both exposure and outcome, the second row represents sample size 1,000 for exposure and sample size 2,000 for outcome, the third row represents sample size 2,000 for exposure and sample size 1,000 for outcome, and the fourth row represents sample size 2,000 for both exposure and outcome. The three columns represent the scenarios where 100%, 80%, and 50% of the variants have positive effects on the exposure. The X axis shows the proportions of causal variants in the signal region, with 15% and 35% causal variants respectively.

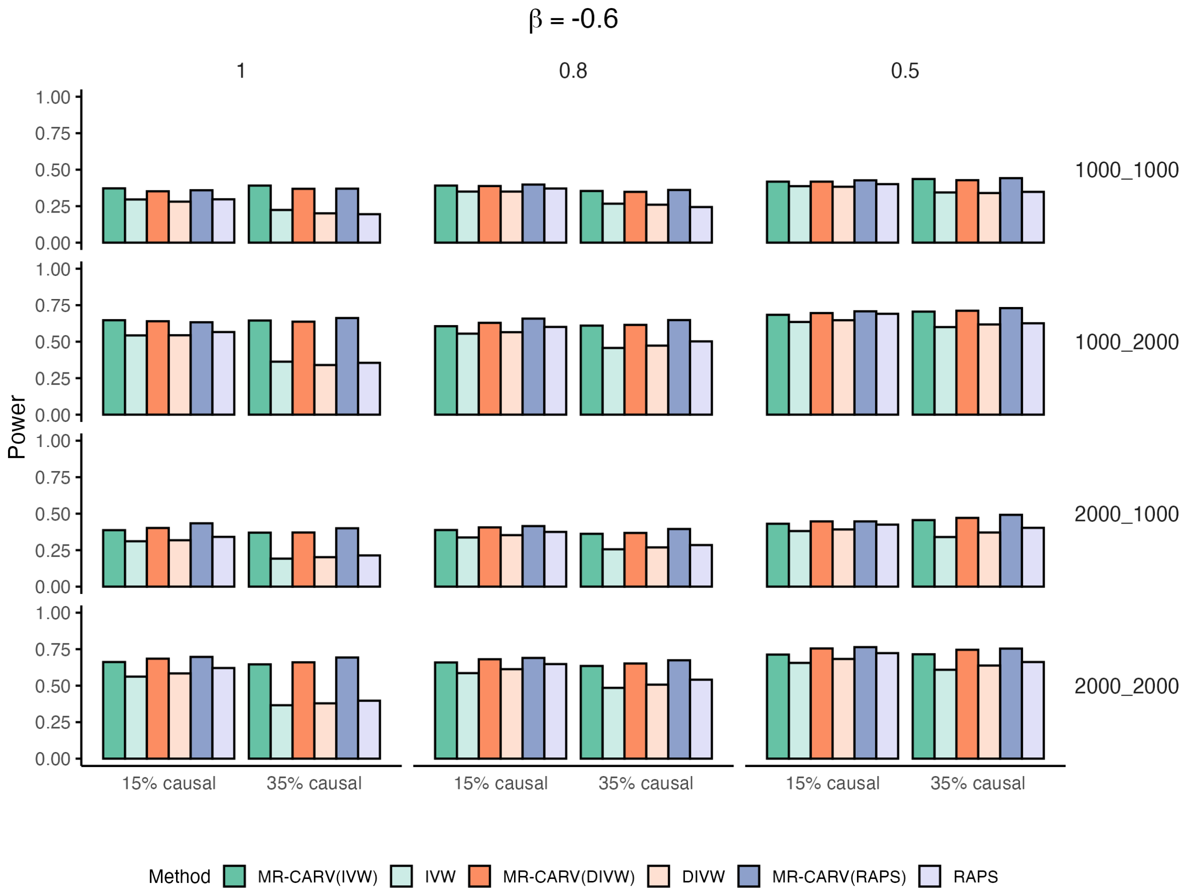


**Figure S9: Power results under 1,000 simulations when $\beta = -0.6$ for binary exposure and continuous outcome.** The first row represents sample size 1,000 for both exposure and outcome, the second row represents sample size 1,000 for exposure and sample size 2,000 for outcome, the third row represents sample size 2,000 for exposure and sample size 1,000 for outcome, and the fourth row represents sample size 2,000 for both exposure and outcome. The three columns represent the scenarios where 100%, 80%, and 50% of the variants have positive effects on the exposure. The X axis shows the proportions of causal variants in the signal region, with 15% and 35% causal variants respectively.

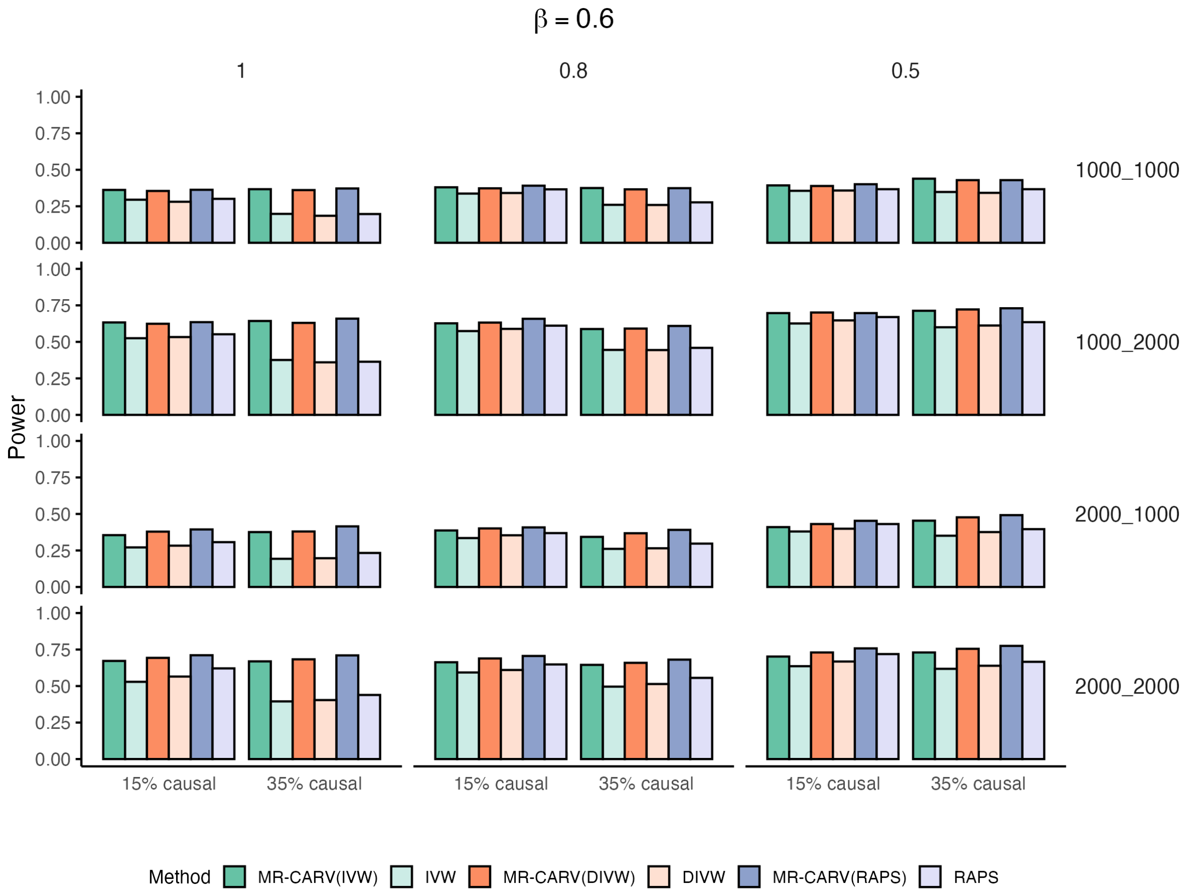


**Figure S10: Power results under 1,000 simulations when $\beta = 0.6$ for binary exposure and continuous outcome.** The first row represents sample size 1,000 for both exposure and outcome, the second row represents sample size 1,000 for exposure and sample size 2,000 for outcome, the third row represents sample size 2,000 for exposure and sample size 1,000 for outcome, and the fourth row represents sample size 2,000 for both exposure and outcome. The three columns represent the scenarios where 100%, 80%, and 50% of the variants have positive effects on the exposure. The X axis shows the proportions of causal variants in the signal region, with 15% and 35% causal variants respectively.

#### 3.4 Binary Exposure, Binary Outcome

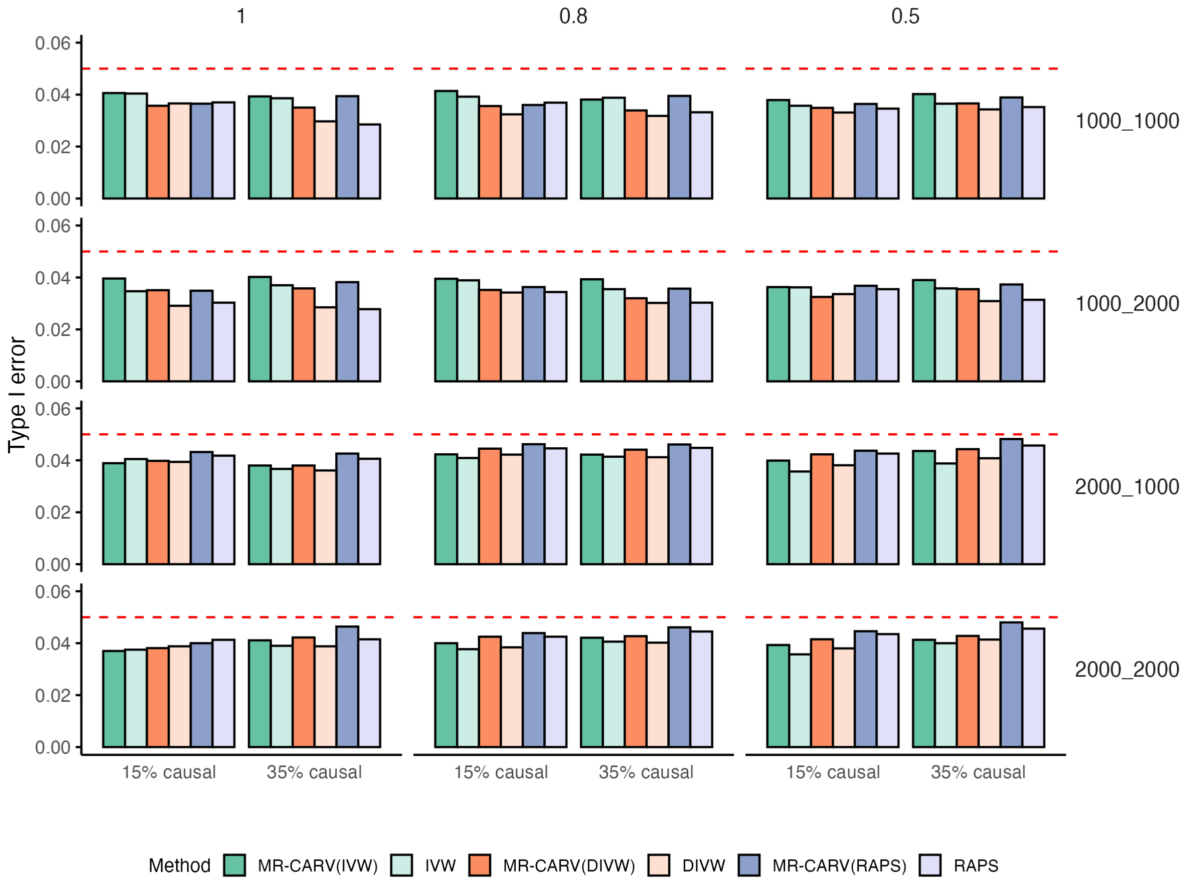


**Figure S11: Type I error rate under 10,000 simulations for binary exposure and binary outcome.** The first row represents sample size 1,000 for both exposure and outcome, the second row represents sample size 1,000 for exposure and sample size 2,000 for outcome, the third row represents sample size 2,000 for exposure and sample size 1,000 for outcome, and the fourth row represents sample size 2,000 for both exposure and outcome. The three columns represent the scenarios where 100%, 80%, and 50% of the variants have positive effects on the exposure. The X axis shows the proportions of causal variants in the signal region, with 15% and 35% causal variants respectively.

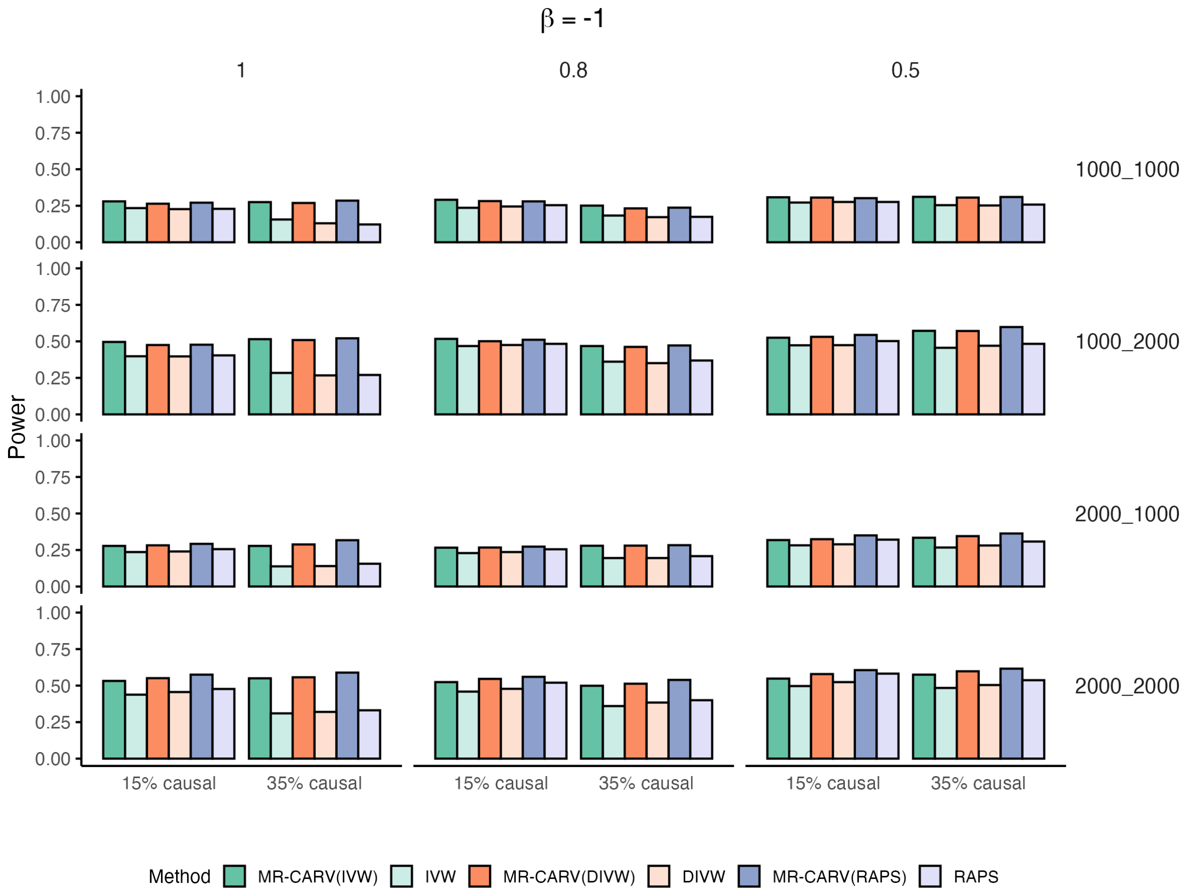


**Figure S12: Power results under 1,000 simulations when $\beta = -1$ for binary exposure and binary outcome.** The first row represents sample size 1,000 for both exposure and outcome, the second row represents sample size 1,000 for exposure and sample size 2,000 for outcome, the third row represents sample size 2,000 for exposure and sample size 1,000 for outcome, and the fourth row represents sample size 2,000 for both exposure and outcome. The three columns represent the scenarios where 100%, 80%, and 50% of the variants have positive effects on the exposure. The X axis shows the proportions of causal variants in the signal region, with 15% and 35% causal variants respectively.

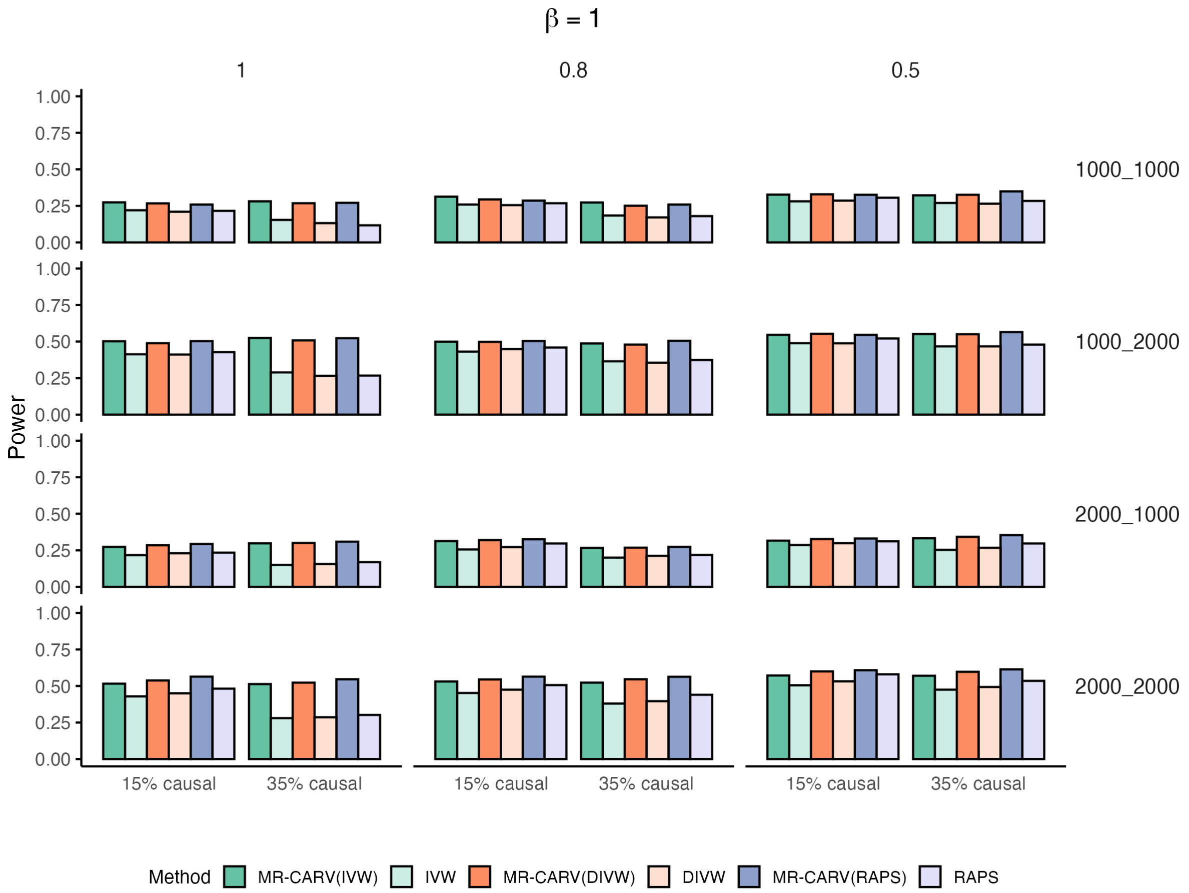


**Figure S13: Power results under 1,000 simulations when $\beta = 1$ for binary exposure and binary outcome.** The first row represents sample size 1,000 for both exposure and outcome, the second row represents sample size 1,000 for exposure and sample size 2,000 for outcome, the third row represents sample size 2,000 for exposure and sample size 1,000 for outcome, and the fourth row represents sample size 2,000 for both exposure and outcome. The three columns represent the scenarios where 100%, 80%, and 50% of the variants have positive effects on the exposure. The X axis shows the proportions of causal variants in the signal region, with 15% and 35% causal variants respectively.

#### 4 Simulation under LD Pruning

To assess the robustness of MR-CARV under realistic instrument selection, we applied graph-based LD pruning to all simulated variants on each chromosome, as described in the main manuscript. Simulations were conducted under a continuous exposure and outcome setting with sample sizes $N_x = N_y = 1{,}000$, as the primary aim was to evaluate the impact of LD pruning rather than trait type. In addition to our main MR-CARV analyses, we evaluated an alternative approach that mimics practical instrument selection: selecting all common variants and the top rare variant (i.e., the variant with the highest MAF) from each rare variant set, followed by graph-based LD pruning of the combined variant set. We then applied existing MR methods (IVW, dIVW, RAPS) to these pruned instruments, denoted as IVW+TopRare, dIVW+TopRare, and RAPS+TopRare in the results.

**Figure S14** presents type I error and power. All methods controlled type I error well below the nominal 0.05 threshold across all simulation settings, confirming valid inference under LD-pruned instruments. MR-CARV consistently achieved the highest power across all scenarios. Methods that included top rare variants outperformed those using only pruned common variants. However, their performance remained below that of MR-CARV, highlighting the additional benefit of annotation-informed rare variant integration.

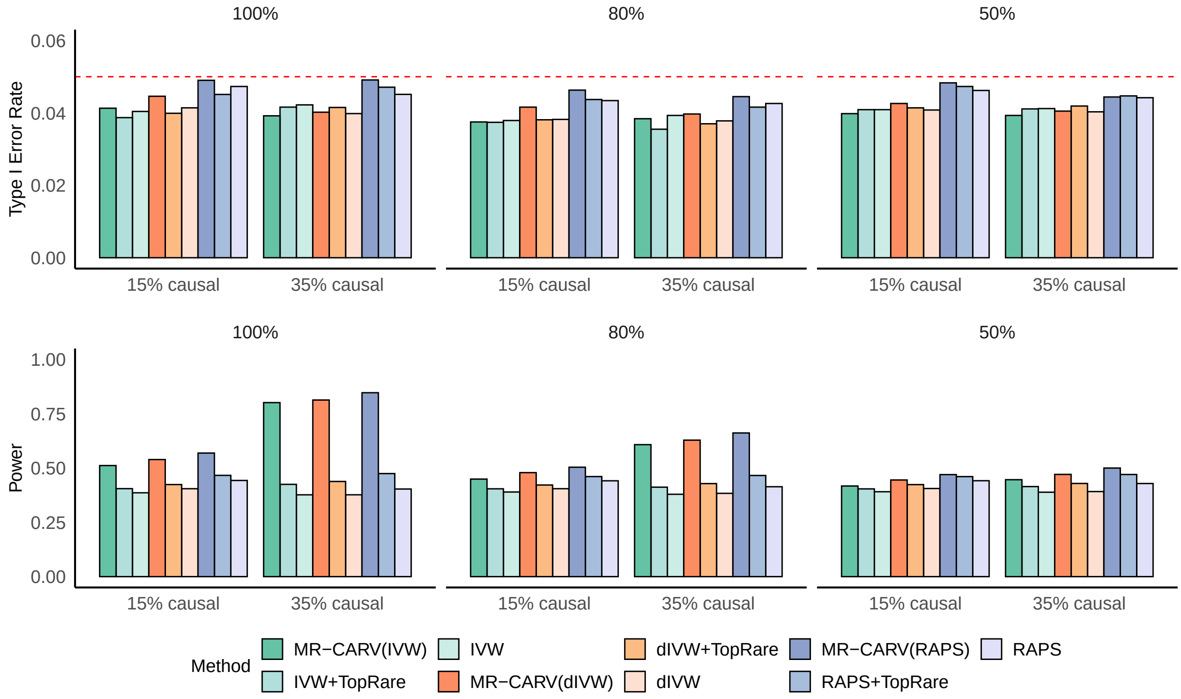


**Figure S14: Type I error rates and power from 10,000 simulations under a continuous exposure and outcome setting with $N_x = N_y = 1{,}000$.** The first row shows type I error rates under the null ($\beta = 0$), and the second row shows power when $\beta = 0.1$. The three columns represent the scenarios where 100%, 80%, and 50% of the causal rare variants have positive effects on the exposure. The X axis shows the proportions of causal rare variants in the signal region, with 15% and 35% causal variants respectively. "+TopRare" indicates the inclusion of the top MAF variant per rare variant set alongside common variants

#### 5 Simulation under Horizontal Pleiotropy

To evaluate the performance of MR-CARV under horizontal pleiotropy, we adopted a simulation framework inspired by Bowden et al. [1], extending it to incorporate rare variants. Unlike the main simulation based on sim1000G data, which was constrained to 8 instruments due to computational overhead, this design allowed a larger number of instruments for evaluating pleiotropy. Specifically, we simulated 25 common variants with MAFs drawn from Uniform$(0.1, 0.5)$, and 25 rare variant sets, each comprising 50 rare variants with MAFs from Uniform$(0.0001, 0.01)$. Sample sizes were set to $N_x = N_y = 1{,}000$. The generative models for the unmeasured confounder $(U)$, exposure $(X)$, and outcome $(Y)$ were as follows:

$$U = \sum_{i=1}^{25} \alpha_i^u G_i + \sum_{j=1}^{25} \sum_{l=1}^{50} \alpha_{jl}^u G_{jl} + \varepsilon_u, \tag{1}$$

$$X = \sum_{i=1}^{25} \alpha_i^x G_i + \sum_{j=1}^{25} \sum_{l=1}^{50} \phi_{jl} \alpha_{jl}^x G_{jl} + U + \varepsilon_x, \tag{2}$$

$$Y = \sum_{i=1}^{25} \alpha_i^y G_i + \sum_{j=1}^{25} \sum_{l=1}^{50} \alpha_{jl}^y G_{jl} + \beta X + U + \varepsilon_y, \tag{3}$$

where $G_i$ and $G_{jl}$ denote the genotypes of the $i$-th common variant and the $l$-th rare variant in the $j$-th set, respectively. Consistent with Bowden et al. [1], instrument strengths of common variants $\alpha_i^x$ were drawn from Uniform$(0.5, 4)$, and error terms $\varepsilon_u, \varepsilon_x, \varepsilon_y$ were sampled from $\mathcal{N}(0, 2)$ distributions. For rare variants, we adopted the same annotation-informed simulation framework as in the main manuscript. The causal status of each rare variant was determined by a binary indicator $\phi_{jl} \in \{0, 1\}$, drawn from a Bernoulli distribution

$$P(\phi_{jl} = 1) = \text{expit}\left(b_0 + \sum_{i=1}^{5} b_i A_i\right),$$

where $A_i \sim \mathcal{N}(0, 1)$ are five randomly selected functional annotations (from a total of ten),

$b_i = \log(5)$, and $b_0$ was set to logit(0.015) or logit(0.18) to yield approximately 15% or 35% causal variant ratios, respectively. For causal variants ($\phi_{jl} = 1$), the effect sizes on exposure was specified as $\alpha^x_{jl} = c_0 \cdot |\log_{10}(\text{MAF}_{jl})|$, with $c_0 \sim \text{Uniform}(0.5, 4)$. To reflect varying levels of directional concordance—relevant for burden-based tests, we set the proportion of positive effects among causal rare variants within each set ($\phi_{jl}\alpha^x_{jl} > 0$) to 100%, 80%, or 50%.

We examined four pleiotropy scenarios. The first three (no pleiotropy, balanced pleiotropy, and directional pleiotropy) satisfy the InSIDE (Instrument Strength Independent of Direct Effect) assumption, while the final one explicitly violates this assumption:

- No pleiotropy, InSIDE satisfied: $\alpha^y_i = 0,\ \alpha^y_{jl} = 0,\ \alpha^u_i = 0,\ \alpha^u_{jl} = 0$
- Balanced pleiotropy, InSIDE satisfied: $\alpha^y_i \sim \mathcal{N}(0, 0.05^2),\ \alpha^y_{jl} \sim \mathcal{N}(0, 0.05^2),\ \alpha^u_i = 0,\ \alpha^u_{jl} = 0$
- Directional pleiotropy, InSIDE satisfied: $\alpha^y_i \sim |\mathcal{N}(0, 0.05^2)|,\ \alpha^y_{jl} \sim |\mathcal{N}(0, 0.05^2)|,\ \alpha^u_i = 0,\ \alpha^u_{jl} = 0$
- Directional pleiotropy, InSIDE violation: $\alpha^y_i \sim |\mathcal{N}(0, 0.05^2)|,\ \alpha^y_{jl} \sim |\mathcal{N}(0, 0.05^2)|,\ \alpha^u_i \sim \text{Uniform}(0, 0.05),\ \alpha^u_{jl} \sim \text{Uniform}(0, 0.05)$.

To evaluate type I error, we set the causal effect of the exposure on the outcome to $\beta = 0$, and to assess power, we used $\beta = 0.02$. Each scenario was repeated across 10,000 simulation replicates, and inference was conducted at a significance level of 0.05. We compared existing MR methods—including IVW (using both first-order and second-order approximations for the variance of the ratio estimate), dIVW, and RAPS—with our proposed MR-CARV framework that incorporate rare variants. To provide a more comprehensive evaluation across variant types, we additionally report results from rare variants alone, offering insight into their independent informativeness under varying pleiotropy structures. Further, we report the average variance in exposure explained ($R^2$) by common variants, rare variants, and both combined, to assess instrument strength across settings. Full results are provided in Supplementary Table S3.

Under scenarios of no pleiotropy or balanced pleiotropy (InSIDE assumption satisfied), all methods maintained appropriate type I error control. However, in settings involving directional pleiotropy or InSIDE violations, all the methods exhibited inflated type I error, consistent with their underlying assumptions. Notably, rare-variant-only analyses consistently showed conservative type I error, particularly when causal variant ratios were low or effect directions were inconsistent (e.g., 50% positive ratio). This behavior may warrant further investigation. In terms of power, when type I error was adequately controlled (e.g., under no or balanced pleiotropy), including both common and rare variants (MR-CARV) led to higher power compared to using either variant class alone. Notably, the gains were more pronounced when the causal ratio and positive ratio among rare variants were higher, reflecting greater variance in exposure explained by rare variant sets.

### References

[1] Bowden, J., Davey Smith, G., and Burgess, S. Mendelian randomization with invalid instruments: effect estimation and bias detection through Egger regression. In: *International journal of epidemiology* 44.2 (2015), pp. 512–525. DOI: https://doi.org/10.1093/ije/dyv080.
